# Evidence-processing errors and their correction in an LLM-assisted systematic review: a retrospective methodological case study

**DOI:** 10.64898/2026.09.21.26363538

**Authors:** Chuan Yin, Zehao Jing, Zhicheng Zhang

## Abstract

**Background:** Errors in interpreting sources, identifying overlapping cohorts and handling corrections can change the results of systematic reviews. We investigated how documented evidence-processing errors affected one completed review assisted by large language models (LLMs), and how an auditable workflow linked their detection and correction to released outputs.

**Methods:** We conducted a retrospective, non-blinded methodological case study of a registered prognostic review. Frozen correction and audit records were reconciled into distinct root-cause events. We described their recorded consequences for analysis membership, study weights, pooled results, certainty and publication files, and traced four illustrative error pathways to implemented controls. Six release-integrity domains provided supporting internal checks. The production review was also the development case; there was no external comparator or independent human reference.

**Results:** Fifty root-cause events were documented; 12 had changed pooled results before correction. Forty-six were resolved and four remained disclosed non-critical limitations. Twenty-one events had at least one recorded scientific consequence. The four pathways concerned survival time origin, cohort dependence, outcome direction and certainty assessment. Corrections changed analysis membership, weighting or interpretation; in the certainty case, the risk-of-bias domain changed while the final certainty grade remained at its lowest level. The application comprised 454 reports and 445 studies, with 41 of 49 registered analyses fitted. Public records link the reported defects, corrections, rebuilds and release checks.

**Conclusions:** In this review, consequential errors arose in the relationships between source judgments and their downstream use. Explicit estimand rules, adjudication-driven dependence mapping, source-based outcome coding and documented downstream rebuilds made corrections inspectable. The case provides operational rules and documented failure examples for other review teams to examine and test; it does not establish error rates or comparative effectiveness.

**Systematic review registration:** Underlying clinical review: PROSPERO CRD420261449181. The retrospective methods evaluation was not prospectively registered.

## Background

Large language models (LLMs) support screening, extraction and synthesis in systematic reviews, but accurate individual outputs do not ensure a valid final synthesis. A correctly transcribed estimate may refer to the wrong outcome, population or time origin. Two accurate extractions may describe overlapping participants, and an obsolete table may remain in circulation after its analysis has been corrected. These problems arise between review stages as well as within individual tasks. Errors in human screening and extraction, and overlap in evidence synthesis, provide an established context for their assessment.[1-3]

Recent systems address multiple review tasks, including review replication and updating.[4-8] Evaluations using human references or published reviews report variable performance across tasks and study designs.[9-18] EviSearch combines structured extraction, cell-level attribution and human auditing; other frameworks emphasize reviewable evidence packages or retrieval with audit feedback.[19-21] These developments support a broader evaluation question: whether source judgments retain their intended meaning and statistical role throughout the production of a review. Human responsibility and transparent disclosure remain necessary when AI is used in evidence synthesis.[22, 23] In review-level evaluations, close approximation of a published pooled estimate and reliable production of a complete review are different outcomes. Pratte et al examined approximation of critical-care meta-analyses, whereas Zou et al found that strong screening performance did not ensure reliable quantitative synthesis at manuscript level.[16, 17] An extraction benchmark also identified omissions as an important error source.[18] These findings motivate inspection of the relationships between extracted fields and their downstream uses, alongside assessment of individual task outputs.

We developed an auditable evidence system during a prognostic review of preoperative social connection and postoperative outcomes. Prognostic reviews are a demanding use case because publications may share patients, an exposure may appear only as a covariate, and follow-up may begin at diagnosis or surgery. These distinctions determine which estimates can answer the review question.[24, 25]

The system linked source interpretation, evidence identities, analysis-specific statistical contributions, correction records and released files. We call the compilation process an evidence compiler and its documented output an accountable evidence release (AER). AER is a project-defined operational specification, not a certification standard. Here, the completed clinical review serves as a real application case for investigating documented processing errors and their correction; it is the same review used to develop the system.

We asked which documented evidence-processing errors changed the synthesis, how those errors were detected and corrected, and which implemented controls made the correction traceable to released outputs. The historical error record and four illustrative pathways formed the primary evaluation; release-integrity checks supplied supporting evidence. The objective was to derive inspectable operational lessons for review teams, rather than to estimate comparative accuracy, reviewer workload or clinical benefit.

## Methods

### Study design and application

We conducted a retrospective, non-blinded methodological case study of one registered prognostic review (PROSPERO CRD420261449181). PRISMA 2020 guided the underlying review.[26] The system and signed release preceded finalization of this evaluation; the analysis plan was hashed after release to support reproducible derivations, rather than prospectively prespecified. All scientific values came from frozen release SR-49523c19b885c87a, without refitting clinical effects. [27] Searches of seven databases identified 33,301 records; 12,902 duplicates were removed, leaving 20,399 records for screening. Of 1,979 reports sought, 252 were not retrieved and 177 were retrieved but not production-eligible, leaving 1,550 for eligibility assessment. The identity reconciliation begins with the 1,979 reports sought; its frozen label “records screened” does not denote the full search set. The companion clinical manuscript reports clinical associations and certainty from this completed application. It shares the development release and is not an independent validation dataset. The full selection flow is reported in Figure 1 of the companion clinical manuscript (doi:10.64898/2026.09.15.26363083). Supporting methods and records are in Additional file 1. Additional file 2 maps PRISMA 2020 reporting items to this methodological case evaluation and distinguishes them from reporting of the underlying clinical review.

**Figure 1.**
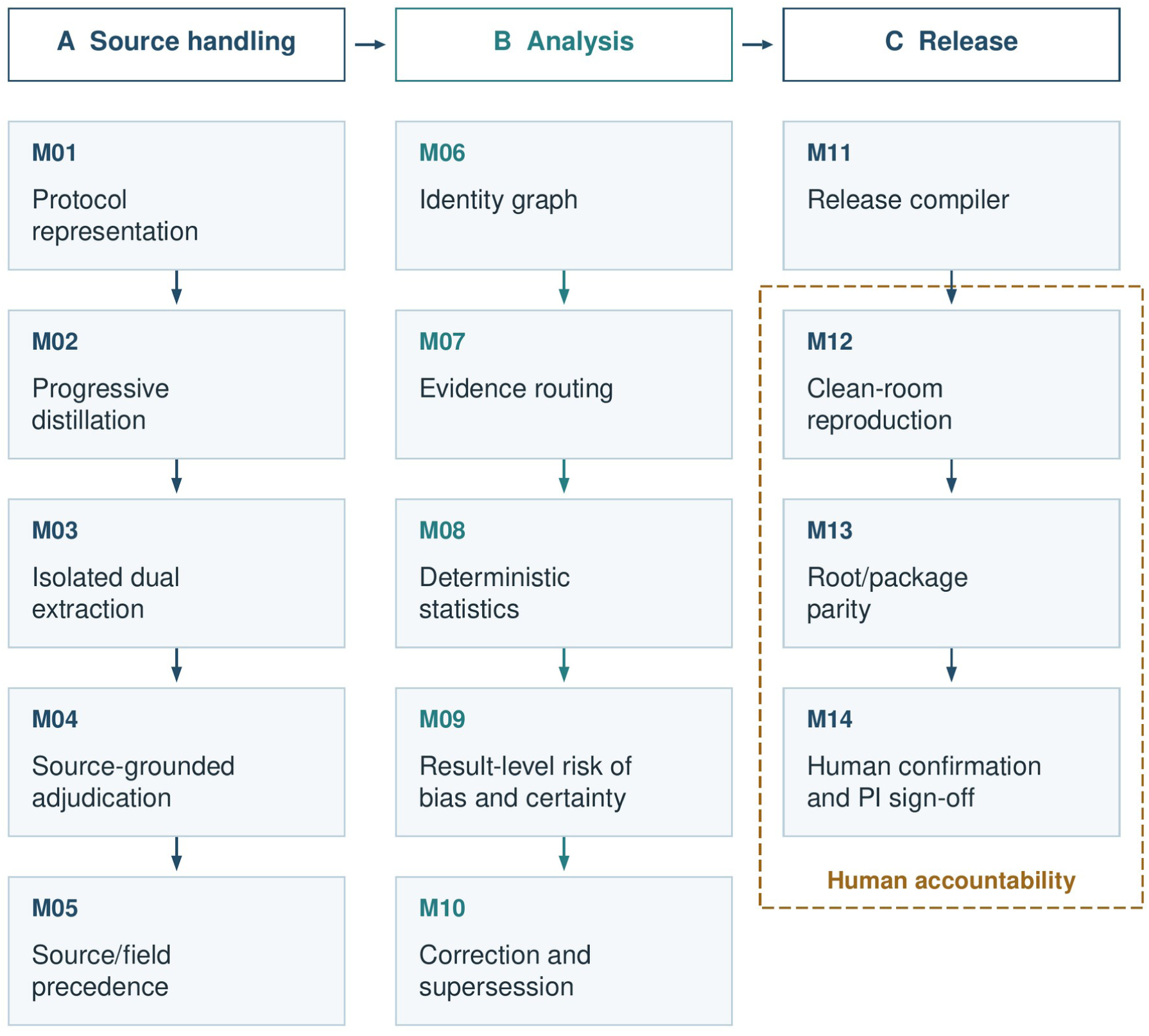
Evidence compiler architecture. Fourteen modules cover source handling (A), analysis (B) and release (C). Arrows indicate processing order. The dashed boundary marks human accountability for final verification and release authorization. Automated checks operate within this boundary, and human involvement also occurs at earlier stages. The principal investigator adjudicates material escalations, freezes decision rules and authorizes release. Automated checks can block release when specified conditions are not met. Clean-room reproduction used a separate implementation within the same project.

### Event identification and classification

We retrospectively counted distinct root-cause events with confirmed terminal statuses from the correction-cycle ledger, verified release-blocking findings from Target 4, and audit-programme status. Repeated records, recurrence across audit cycles and downstream stale files caused by the same defect were not separate events. Sibling defects were merged when they shared a root cause. Unconfirmed allegations and findings that did not satisfy the release-blocking criterion were excluded, with their crosswalk retained. Consequences could overlap across analysis membership, k, pooled results, certainty, denominators and manuscript packages. Terminal statuses distinguished completed downstream rebuilds, verified fixes and disclosed non-critical limitations without fixes. These descriptive counts characterize documented history, not error incidence or errors prevented (Supplementary Table S6).

The event total came from 63 correction-ledger rows: removing 17 repeated rows left 46 defect records, then merging sibling records by shared root cause yielded 39 events. Ten verified release-blocking Target 4 findings and one audit-infrastructure event completed the total of 50. The event crosswalk preserves these transformations, including identifiers containing more than one defect record (Supplementary Table S6). Four illustrative error pathways were selected to show distinct mechanisms, not as a representative sample or a prospective challenge set. Historical before-and- after point estimates were reported only when the correction ledger explicitly recorded both values and the effect type could be verified against the analysis registry. These snapshots were distinguished from final clinical outputs; paired confidence intervals and effects attributable to an isolated correction were not inferred.

### Evidence processing and correction

The system comprised 14 modules spanning source handling, analysis and release (Figure 1; Supplementary Table S1). LLM agents supported screening, extraction, proposed source adjudications, code development, coordination and audits. Machine-readable contracts specified admissible evidence and synthesis conditions; deterministic code performed statistical calculations and release checks. Investigators resolved material ambiguities and authorized release. Two extraction passes were isolated from one another, with a third pass adjudicating disagreements against printed source text. Majority agreement alone could not resolve a discrepancy. Fields retained an ordered source-authority state, from adjudicated or verified values through extracted and normalized values to detector-derived or unknown values. Lower-authority values could not overwrite higher-authority values, and unknown values remained explicit. Supplementary Table S2 details automated and human work. Evidence moved through six stages, D0–D5, from records to estimates. Information could be discarded only when the rules for later tasks established that it was no longer required.

The identity graph distinguished records, reports, studies, cohort entities, dependence clusters, analysis weight units and estimates (Table 1; Figure 2). A report is a publication; a study may be described in several reports. Cohort entities represent source-defined cohorts or subsets.

**Figure 2.**
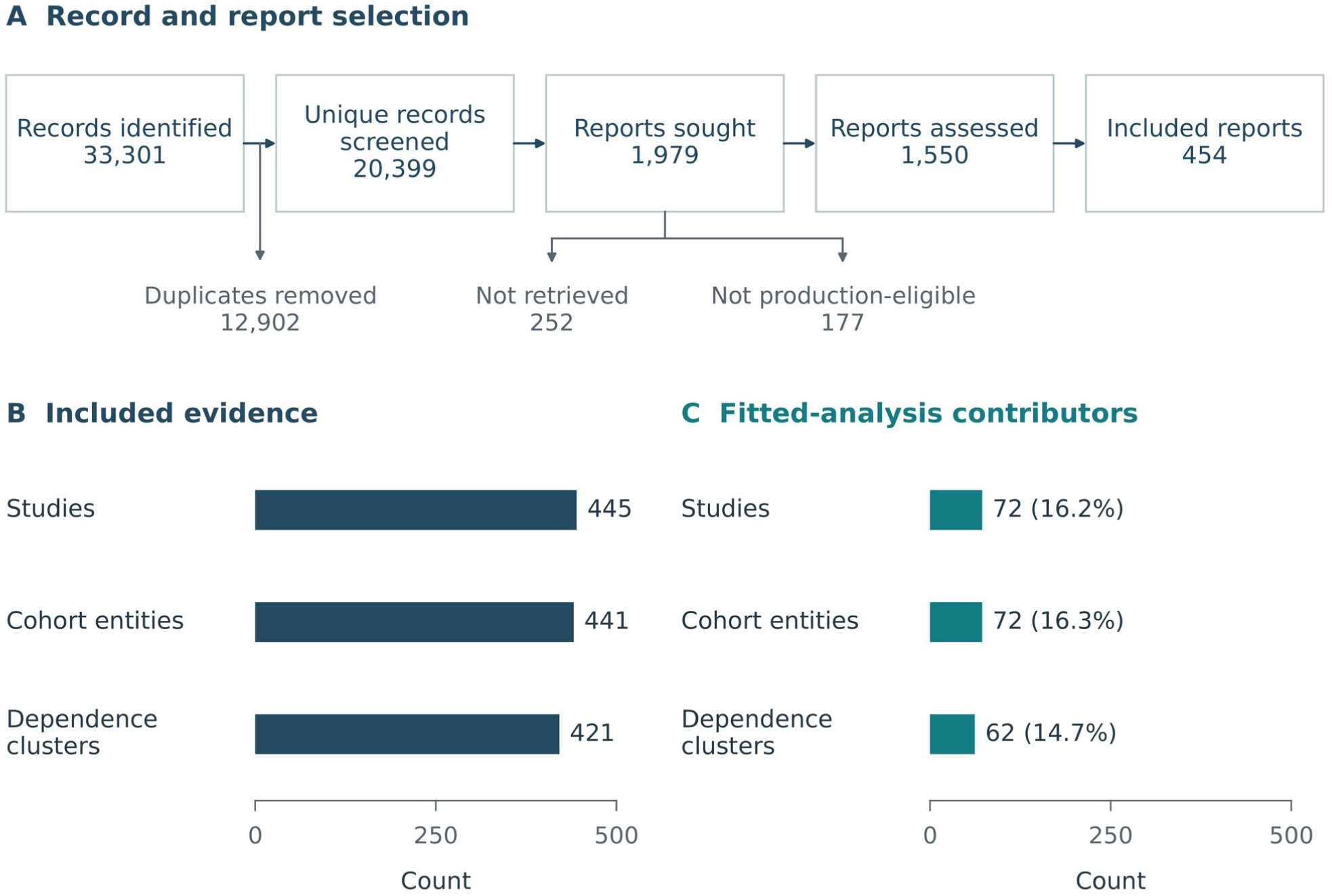
Selection, identity grouping and participation in fitted analyses. (A) Record and report selection. The identity graph begins with reports sought for retrieval. This overview is not a complete PRISMA flow diagram. (B) The 454 included reports map to 445 studies, 441 cohort entities and 421 dependence clusters. These are different counting units, not sequential exclusions. (C) Contributors to fitted analyses, using the same count scale as panel B. Percentages use the corresponding included-evidence denominator: 72/445, 72/441 and 62/421. The 41 fitted analyses comprise 216 analysis-specific weight units and 113 estimates. A cluster can contribute one weight in each of several analyses, and an estimate can serve more than one weight unit.

**Table 1.** Corpus counts and evidence routes

| Quantity | Counting unit | n | Interpretation |
| --- | --- | --- | --- |
| Reports sought for retrieval | RecordID/ReportID | **1979** | Entry point to the identity graph, labelled “records screened” in the release. Follows screening of 20399 unique records (33301 identified). |
| Included reports | ReportID | **454** | Publications |
| Included studies | StudyID | **445** | Multiple reports of the same study counted once |
| Source-defined cohort entities | CohortEntityID | **441** | Reports from the same cohort counted once |
| Dependence clusters | DependenceClusterID | **421** | Shared participants, or overlap cannot be excluded |
| Studies in $\geq 1$ fitted analysis | StudyID | **72** | Studies routed to pooled meta-analysis |
| Cohort entities in $\geq 1$ fitted analysis | CohortEntityID | **72** | |
| Dependence clusters in $\geq 1$ fitted analysis | DependenceClusterID | **62** | |
| Analysis weight units across fitted analyses | AnalysisWeightUnitID | **216** | Cluster contributions specific to each analysis. This is neither a count of independent clusters nor a further selection step. |
| Active estimates across fitted analyses | EstimateID | **113** | Unique estimates across analyses; an estimate can serve several weight units |
| Fitted analyses | MetaAnalysisID | **41 of 49** | 7 not fittable ($k < 2$); 1 not poolable |
| Route: pooled meta-analysis | StudyID | **72 (16.2%)** |  |
| Route: replication target ($k < 2$) | StudyID | **79 (17.8%)** | |
| Route: not poolable | StudyID | **3 (0.7%)** | Incommensurable units |
| Route: no current quantitative synthesis | StudyID | **291 (65.4%)** | Mapped as EVIDENCE_GAP; not evidence of absence |
| Clusters without a terminal quantitative route | DependenceClusterID | **323 of 421 (76.7%)** | Cluster denominator; 98 clusters in directional synthesis |
| Principal-family source records | EstimateID | **39** | Sum of k across five principal families (9+8+9+9+4) |

Upstream PRISMA counts for the underlying review: 252 reports not retrieved; 177 retrieved but not production-eligible; 1,550 assessed.

Dependence clusters group entities sharing participants or with overlap that cannot be excluded. An analysis weight unit is the object permitted to receive one weight in a particular analysis. For each fitted analysis, the selected primary-estimate count and the unique analysis-weight-unit count had to equal the reported k (the number of analysis weight units in that analysis). A hierarchy selected estimates using primacy, exposure focality, cohort coverage, outcome and time, adjustment, completeness and source authority. Selection could not depend on statistical significance or row order; a hierarchy level could distinguish candidates only when all were assessable at that level. Admissible alternatives entered a multiverse analysis.[28]

Each study received a terminal route: pooled meta-analysis, replication target with k<2, not poolable because of incompatible units, or no current quantitative synthesis route (Supplementary Figure S3). The internal code EVIDENCE_GAP denotes the last state under the frozen contracts; it does not establish absence of an association or a global evidence gap. Analyses with insufficient or incompatible inputs retained explicit non-fitted states. Effect measures remained separate. Random-effects models used restricted maximum likelihood and modified Hartung–Knapp intervals whose variance factor could widen, but not narrow, conventional random-effects intervals.[29-31] Prespecified thresholds permitted prediction intervals at k≥5 and small-study-effect tests at k≥10; otherwise, an explicit not-reported code was retained.[32-34] DerSimonian–Laird estimation supplied a starting value and historical reference.[35] Result-level assessments used the Quality In Prognosis Studies (QUIPS) tool.[36] Formal principal-family certainty followed Grading of Recommendations Assessment, Development and Evaluation (GRADE) guidance for prognostic factors.[37] Mapping, prognostic association, magnitude, transportability, individual applicability and causal-intervention claims remained separate.

Ten release properties covered source traceability, identity, dependence weighting, estimand compatibility, deterministic statistics, terminal states, correction propagation, supersession, release-to-package parity and human authorization (Table 2). Correction records linked affected objects, root causes, consequences, preventive controls and downstream rebuild status. Derived artifacts were regenerated from frozen inputs, while superseded or invalidated files were retained as history and blocked from publication packages. Targets were hashed before audit, and generation was prohibited while a freeze marker was active. Four unsuccessful attempts at autonomous fixed-target certification were retained as diagnostic archives and supplied no final scientific estimate. The final release was explicitly supervised and authorized by the principal investigator (PI); its manifest prohibits the description “fully autonomous clean release”. Once a correction was confirmed, its record specified whether dependent membership files, statistical results, certainty assessments and publication outputs required regeneration. Human escalation records retained the competing options and their measured consequences. The PI resolved these escalations, froze statistical and certainty rules and signed a release with a defined scope, rather than authorizing an unrestricted autonomous pipeline.

**Table 2.** Design properties of an accountable evidence release

| Property | Artifact | Check | Release evidence | Scope | Limitation |
| --- | --- | --- | --- | --- | --- |
| Source traceability | Source locator per analysed estimate; bounded source audit | Source-locator components checked against the source; terminal source state | D1 39/39; chain audit 39/39 | Principal families; locators for all analysed estimates | Non-principal locators checked mechanically where possible; not re-traced here |
| Identity | Record → report → study → cohort reconciliation | Unique identifiers at each level; multi-report collapse | 454 reports → 445 studies → 441 cohort entities | All included studies | Identity adjudicated under rules and by the PI; no external validation |
| Dependence weighting | Clusters, analysis weight units and membership table | One weight per cluster per analysis; count equalities | D2 5/5; D3 5/5 | Invariants: five principal families; membership: all analyses | Invariant holds by construction; unresolved overlap treated conservatively |
| Estimand compatibility | Synthesis contracts for each family; vocabularies for time origin and outcome | Pooling keys: outcome, measure, time origin, contrast | Survival nodes by origin; core/full readmission sets | Registered analyses | Contracts encode project decisions; mismatches not encoded in the contracts cannot be detected |
| Deterministic statistics | Version-locked REML + modified Hartung–Knapp; prediction-interval and small-study-test thresholds | Separate implementation within the same project | D4 1217/1217 | 41 fitted analyses | Reproduced within the project; both implementations may share specification assumptions |
| Refusal and terminal states | Study routes; not-fittable and not-poolable rows | Exactly one route per included study | 445 routes; 7 not fittable; 1 not poolable | All included studies and registered analyses | Complete routing does not establish that each decision not to synthesize is correct |
| Correction propagation | Event ledger: consequences and rebuild status | Object, root cause, consequence, control and status per event | 50 events; 46 closed; 4 disclosed | Final correction cycle and Target 4 triage | Retrospective; unrecorded corrections cannot be checked |
| Supersession | Manifest with status per artifact | Exclusive status; block superseded/invalidated artifacts | 18 superseded, 2 invalidated of 51 | Release artifacts | Earlier manifest had contradictory status (FE-42) and marked a shipped table as superseded (FE-45) |
| Root-to-output parity | Release manifest; SHA-256 comparison; content checks | Byte identity and content consistency | D5 94/94; 83/83 checks | Two manuscript packages | Byte identity does not establish semantic validity. Tables derived for this manuscript are outside the ledger. |
| Human authorization | Hashed PI sign-off; two-person registry; escalation cards | Archived signature; record-level registry | D6 39/39; signed 2026-08-31 | Principal-family records; release decisions | Recommendation-assisted, not blinded; separate-agent audit A incomplete |

These properties are an operational specification, not scored experiments or a certification standard.

### Supporting release checks

Six supporting release-integrity domains were reported separately (Table 3): terminal source states, one weight per dependence cluster, equality of selected-estimate counts, unique weight-unit counts and reported k, same-project numerical reproduction, byte identity between release and publication files, and two-person confirmation. These domains overlap with the ten design properties and are not independent experiments or a composite score. Dependence checks assess conformance to the encoded graph, not the correctness of every overlap judgment. The numerical comparison used two implementations developed within the project; byte comparisons establish file identity. Neither is an external validation standard. A deposited machine-readable AER instance records the contracts, sources, identities, decisions, corrections, checks, limitations and authorization. Selected public conclusions were re-traced; this did not establish traceability for every conclusion.

**Table 3.**
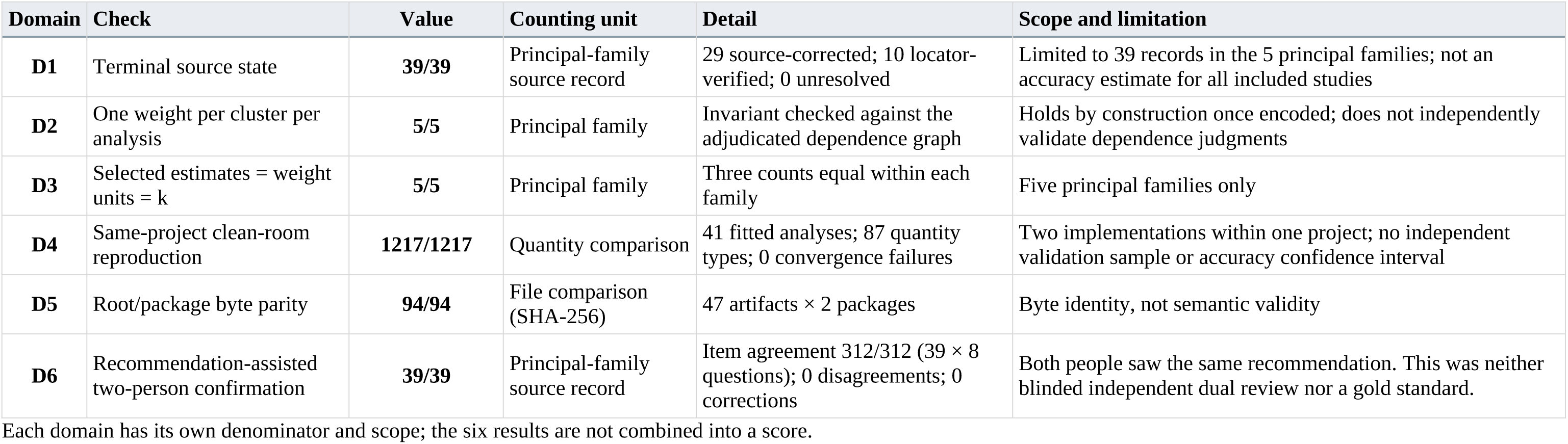
Six release-integrity domains

Chuan Yin and Zehao Jing separately reviewed 39 source records in five principal analysis families, using different deterministic row orders and without seeing each other’s answers. Both saw the same source-grounded recommendations. Eight questions covered identity, exposure and comparator, outcome and time, numerical effect, transformation, model selection, dependence and membership, followed by a final decision. The console permitted an initial judgment before expanding the rationale; the PI used this option for two records and the second reviewer for none. The procedure was therefore recommendation-assisted confirmation, rather than an independent human reference. The PI adjudicated material escalations and signed the release. R1 derivations regenerate evaluation outputs from the frozen release and were executed during manuscript preparation; deposited R2 engines permit statistical refitting but were not rerun. Exact R3 repetition of the agent pipeline is unavailable because runtime builds were not retained and outputs are nondeterministic.

## Results

Fifty distinct root-cause events were documented, including 12 that had changed a pooled result before correction (Figure 3; Table 4). Thirty-nine events were closed with downstream rebuilds completed and seven as fixed and verified; four remained disclosed non-critical limitations.

**Figure 3.**
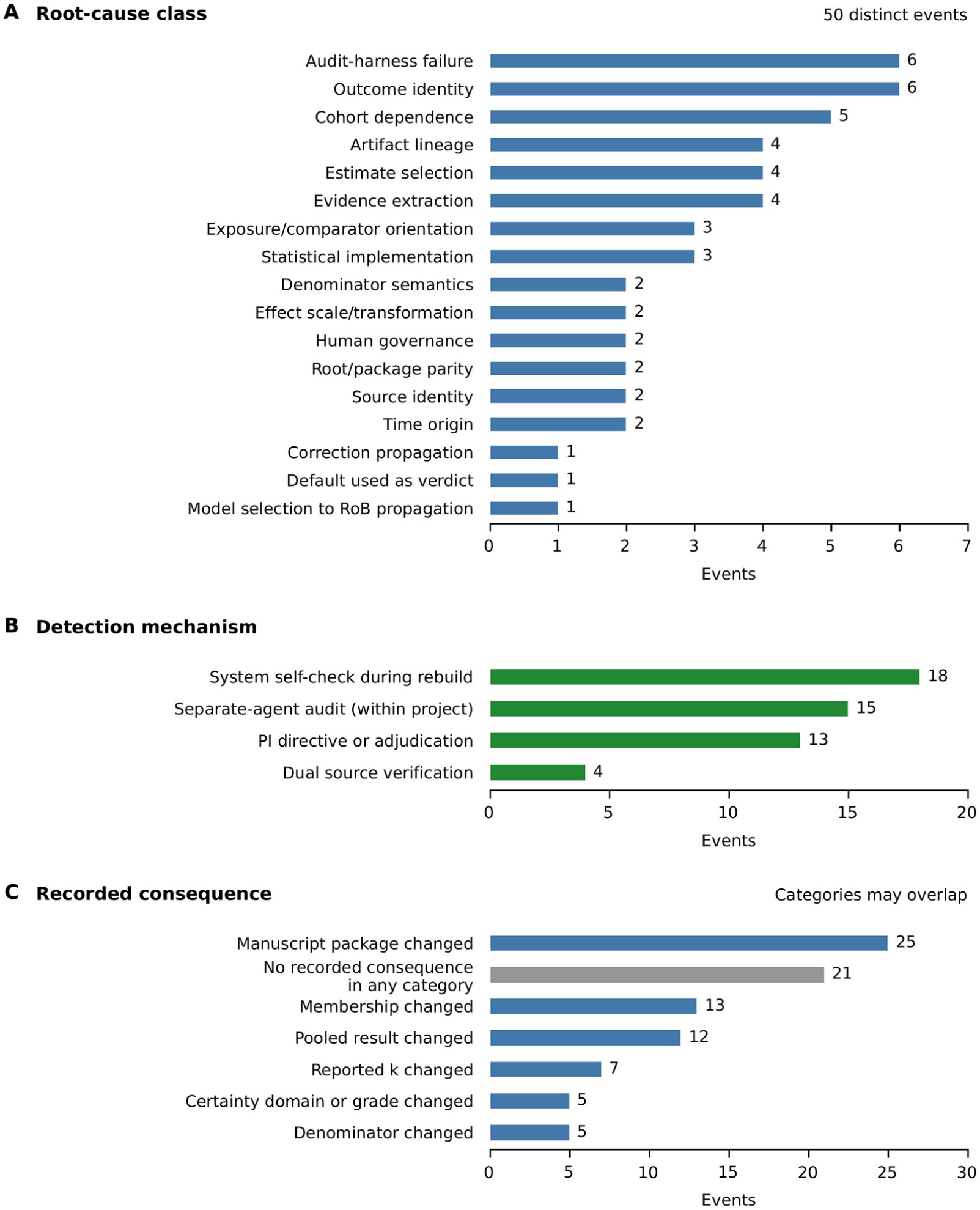
Failure events and recorded consequences. Fifty historical events are classified by root cause (A), recorded detection mechanism (B) and consequence (C). Root-cause and detection categories each account for all 50 events. Consequence categories overlap, and the horizontal-axis range differs between panels. The 21 events with no recorded consequence in any category differ from the 29 without a scientific consequence, which also include 8 with manuscript- package changes only. Separate-agent audits took place within the project. Dual source verification used A/B passes and third-pass adjudication, rather than final human confirmation. The recorded detection category does not establish when an event was first identified. These counts are descriptive, not estimates of an error rate. PI, principal investigator. RoB, risk of bias. k, number of analysis weight units within an analysis.

**Table 4.**
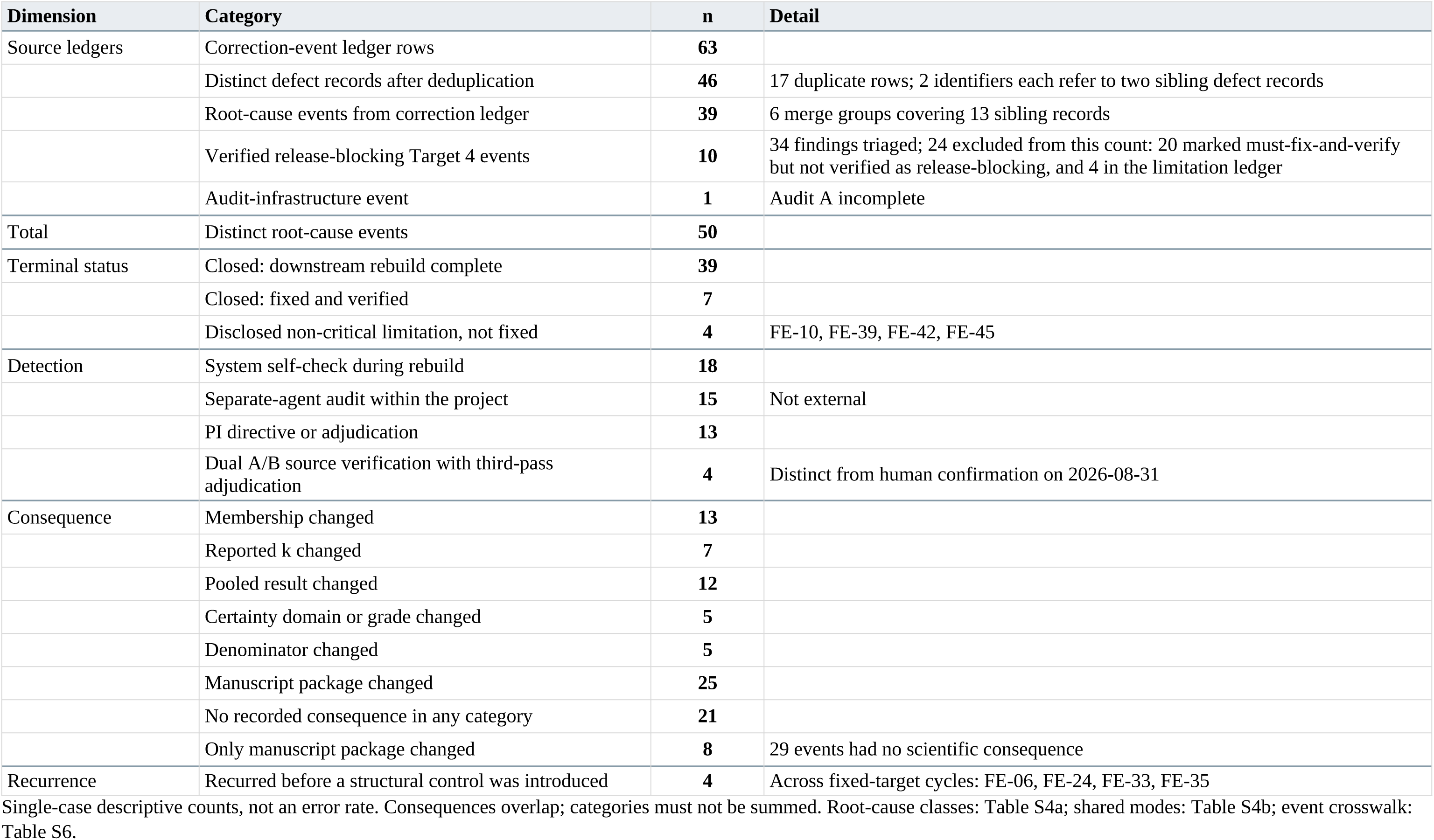
Documented failure events

| Dimension | Category | n | Detail |
| --- | --- | --- | --- |
| Source ledgers | Correction-event ledger rows | 63 |  |
|  | Distinct defect records after deduplication | 46 | 17 duplicate rows; 2 identifiers each refer to two sibling defect records |
|  | Root-cause events from correction ledger | 39 | 6 merge groups covering 13 sibling records |
|  | Verified release-blocking Target 4 events | 10 | 34 findings triaged; 24 excluded from this count: 20 marked must-fix-and-verify but not verified as release-blocking, and 4 in the limitation ledger |
|  | Audit-infrastructure event | 1 | Audit A incomplete |
| Total | Distinct root-cause events | 50 |  |
| Terminal status | Closed: downstream rebuild complete | 39 |  |
|  | Closed: fixed and verified | 7 |  |
|  | Disclosed non-critical limitation, not fixed | 4 | FE-10, FE-39, FE-42, FE-45 |
| Detection | System self-check during rebuild | 18 |  |
|  | Separate-agent audit within the project | 15 | Not external |
|  | PI directive or adjudication | 13 |  |
|  | Dual A/B source verification with third-pass adjudication | 4 | Distinct from human confirmation on 2026-08-31 |
| Consequence | Membership changed | 13 |  |
|  | Reported k changed | 7 |  |
|  | Pooled result changed | 12 |  |
|  | Certainty domain or grade changed | 5 |  |
|  | Denominator changed | 5 |  |
|  | Manuscript package changed | 25 |  |
|  | No recorded consequence in any category | 21 |  |
|  | Only manuscript package changed | 8 | 29 events had no scientific consequence |
| Recurrence | Recurred before a structural control was introduced | 4 | Across fixed-target cycles: FE-06, FE-24, FE-33, FE-35 |

Recorded consequences overlapped: 13 events changed membership, seven changed k, 12 a pooled result, five a certainty domain or grade, five a denominator and 25 a manuscript package. Twenty- one had at least one scientific consequence; eight changed only the package and 21 had no recorded consequence. Thus, 29 had no recorded scientific consequence. These counts include code, integration, audit and governance defects and cannot be attributed entirely to LLM outputs.

The largest root-cause classes were audit-harness failure and outcome identity (six each), cohort dependence (five), and artifact lineage, estimate selection and evidence extraction (four each). Recorded detection routes were self-checks during rebuilds (18), separate-agent audits within the project (15), PI directives or adjudication (13), and dual source-verification passes with third-pass adjudication (four). These categories do not establish first detection. Four events recurred across fixed-target cycles before a structural preventive control was added. Table 5 links four illustrative cases to their detection, correction consequences and implemented controls; full records appear in Supplementary Table S5.

**Table 5.** Error pathways, correction consequences and implemented controls

| Event and error entry | Detection and correction | Recorded consequences | Reusable rule and record |
| --- | --- | --- | --- |
| FE-21: survival time origin. Diagnosis-origin estimates carried most of the weight in a postoperative-survival family. | Target 4 internal audit P05-01 identified the time-origin problem. Origin-specific estimands were separated: postoperative survival became primary and all-origin survival a sensitivity analysis. Related FE-20 restricted detection to explicit source statements. | Membership, k, pooled result, certainty and publication package changed. The recorded history supports a change in the synthesis, without a complete paired effect estimate reported here. | Specify follow-up origin in the analysis contract and use the matching estimand. FE-21: S5 Panel A; S6, Target 4 finding P05-01 (BOUNDED_RELEASE_TARGET4_FINDING_TRIAG E.csv). FE-20 is a separate related correction. |
| FE-06: cohort dependence. A correct overlap adjudication did not enter the manually assembled graph; duplicate weight survived the first correction. | Target 1 audit P07-01 identified the defect; it recurred in Target 2. The enumerator was changed to read the adjudication ledger directly and check for missing declared members. Closure followed the downstream rebuild. | Membership, k, pooled result, denominators and publication package changed. The correction altered which units received weight, rather than simply changing an explanatory label. | Generate dependence links from structured adjudication and fail the check when declared members are absent. FE-06: S5 Panel A; S6, correction-ledger row 22 retained and row 41 duplicate excluded (CE-877R-046). |
| FE-11: outcome direction. Matching the full string allowed “vs skilled nursing facility” to determine the polarity of “discharge home”. | Full-text verification passes A/B, third-pass review and PI patch PI-877R-FINAL-PATCH-01 led to counted-arm matching, comparator stripping, complement masking and corrected desirable/adverse ordering. The higher-is-worse fallback was prohibited. | Membership, pooled result and publication package changed. The recorded detection category was PI directive or adjudication. | Code the event being counted, not the comparator clause or a generic fallback. FE-11: S5 Panel A; S6, ledger rows 39 and 58, two defect records sharing CE-877R-024 and merged as one event. |
| FE-38: model selection and certainty. Corrected results exceeded the protocol’s high-risk-of-bias weight threshold; no quality-restricted set could be constructed. | Live recomputation, documented in correction-cycle section 10, identified the changed risk-of-bias tier. The domain judgment and package were updated without relaxing the protocol criteria. | The certainty-domain judgment and publication package changed. The final GRADE category was already at its lowest level and did not change. | Propagate corrected model selection into certainty records; distinguish domain changes from the final grade. This was a recorded correction, not a newly validated automatic blocker. FE-38: S5 Panel B; S6, ledger row 18 (CE-877R-018). |

### Error pathways and correction consequences

Time origin (FE-21): diagnosis-origin estimates carried most of the weight in a postoperative- survival family. Separating origin-specific estimands changed membership, k, the pooled result, certainty and the package. Postoperative survival became the primary analysis, with all-origin survival retained as a sensitivity analysis. A related correction (FE-20) restricted the time-origin detector to explicit source statements and limited downstream changes.

Cohort dependence (FE-06): an overlap adjudication required two studies to count as one, but manually entered links omitted their cluster and allowed duplicate weight. The defect survived an initial correction. An enumerator was changed to read the adjudication ledger directly, with a check for missing declared members. Correction changed membership, k, the pooled result, denominators and the package. A missing member declared in the overlap ledger now causes the graph check to fail. The event was closed only after the downstream rebuild was completed. For the all-cause-mortality analysis S-OR-1, the correction ledger recorded k changing from 17 to 16 and the pooled OR from 1.2743 to 1.2763.

Outcome direction (FE-11): matching the entire outcome string allowed “vs skilled nursing facility” to determine the polarity of “discharge home”. Verification and PI adjudication led to rules using only the counted arm, removing comparator clauses and masking complement forms before matching. Correction changed membership, the pooled result and the package. Its recorded detection category is PI directive or adjudication. The revised rules also prohibited a higher-is-worse fallback and corrected the ordering of desirable-event and adverse-term handling. At one historical polarity-correction step for the non-home-discharge analysis F3, the ledger recorded a pooled OR change from 1.0663 to 1.3709, with k unchanged. These are correction-stage values, not final released estimates.

Model selection and certainty (FE-38): after correction, high-risk-of-bias results exceeded the protocol’s weight threshold and a quality-restricted set could not be constructed. The risk-of-bias domain moved to a more serious tier. The final certainty grade was already at the lowest level and remained unchanged; the domain and package changed.

### Application and supporting release checks

The corpus comprised 454 reports, 445 studies, 441 cohort entities and 421 dependence clusters (Figure 2; Table 1). Forty-one of 49 registered analyses were fitted, using 216 analysis-specific weight units drawn from 113 distinct estimates across 72 studies, 72 cohort entities and 62 clusters. These units are not 216 independent clusters. All 445 studies reached a terminal route: 72 (16.2%) entered pooled meta-analysis, 79 (17.8%) were replication targets, three (0.7%) had incompatible units and 291 (65.4%) had no current quantitative synthesis route (Supplementary Figure S3). At cluster level, 323 of 421 (76.7%) had no terminal quantitative route; 36 entered direction-only synthesis and 62 also entered effect-size synthesis. Seven registered analyses had k<2 and one had incompatible continuous units, giving eight non-fitted states.

All 39 source records in the five principal families reached terminal states: 29 were corrected against sources and ten verified against structured locators, with none unresolved. Adjudicated values were consistent through the ledger, membership and engine input for all 39. Selected- estimate counts, unique weight-unit counts and reported k agreed in all five families, as did the one-weight-per-cluster check against the encoded graph. Of 81 selected results assessed for bias, 56 were high risk and 25 moderate in the primary aggregation. The certainty registry contained 56 claims across eight types, including five formal prognostic grades. Causal-intervention claims remained not directly evaluated. Supplementary Table S3 identifies the principal analyses and source records, membership and count checks, and release provenance. The complete clinical estimates are reported in the companion manuscript (doi:10.64898/2026.09.15.26363083) and the frozen archive.[27]

For the supporting internal checks, the second implementation reproduced 1,217 of 1,217 numerical comparisons across 41 fitted analyses within prespecified tolerances, covering 87 quantity types, including all 35 principal-family quantities. There were no convergence failures or unexplained disagreements. All 94 file comparisons matched (47 artifacts in two packages); 49 engine-status rows matched the registry, 83 derived-content checks passed and nine final release gates passed.

The final supersession manifest listed 51 artifacts, including 18 superseded and two invalidated artifacts, none distributed under those final statuses. Earlier lineage discrepancies remained disclosed: an earlier manifest assigned conflicting statuses to one artifact (FE-42), and both packages contained a directional-synthesis table identical to one marked superseded in that earlier version (FE-45). FE-44 was closed after adding a check against superseded inputs; distribution had occurred before that correction. These results establish consistency with the signed build within the reported internal scope. Supplementary Figure S1 presents production-scale and release-integrity metrics separately, each with its own denominator.

Both reviewers confirmed all 39 principal records, agreeing on all 312 field-level questions, with no final disagreements or subsequent scientific corrections. This measures agreement after shared recommendations, not independent accuracy. Eight non-critical release limitations remained: three diagnostic-only and five involving supporting artifacts; only four correspond to counted failure events (Supplementary Table S7). Three of four planned internal separate-agent audits completed. Infrastructure failure prevented completion of the main-source audit, which was replaced by deterministic local checks. Source-fragment matching covered 23 of 39 records; nine remaining locators were not mechanically checkable and seven contained no quotable fragment. Their terminal states rely on adjudication and confirmation. A status file within the signed packages also retains a pending decision subsequently resolved in the frozen cluster adjudication: S00498 carries the registry-readmission contribution and S00253 remains linked without weight. The read-only release was retained, with this discrepancy disclosed.

## Discussion

Twelve of the 50 documented root-cause events had changed pooled results before correction. The four detailed pathways show why accurate transcription alone cannot protect a synthesis: an estimate can retain the wrong time origin, an overlap decision can fail to govern weighting, an outcome label can reverse interpretation, and model selection can change a certainty-domain judgment. The clinical review provides a completed application in which these defects and their consequences can be followed through the production records. It supplies evidence about error mechanisms within this case, rather than a comparison of competing workflows.

The cases suggest concrete checks at the transitions between review tasks (Table 5; Supplementary Table S11). Specify the time origin before pooling; generate dependence links from the adjudication ledger and fail the check when declared members are missing; code outcome polarity from the counted event rather than its comparator; and rebuild certainty assessments and publication files after an upstream correction. These rules were implemented in this project. Their preventive effectiveness was not tested independently. Public correction identifiers and artifact records make the reported implementation inspectable. Source verification, numerical reproduction and file comparisons address different questions and must retain separate denominators: a calculation can reproduce an erroneous input, and a byte-identical file can preserve a wrong interpretation.

These connections complement existing systems and evaluation approaches. TrialMind and otto-SR examine several review tasks, with human or review-level comparisons and, for otto-SR, the effects of corrected inputs on pooled results.[4, 7] EviSearch supports source attribution and human auditing, while proposed evidence-package frameworks include reviewer records, change control and source updates.[19, 20] Provenance knowledge graphs and executable paper packages offer related approaches to inspection and reproducibility.[38, 39] These are precedents, not capabilities claimed to be unique to our system. Supplementary Table S9 compares the assessed versions and evaluation conditions. Our emphasis is the implemented linkage between source judgments, analysis-specific weights, recorded correction consequences and the released files within one review. Existing component studies cover active-learning screening, machine-assisted prioritization and structured extraction.[13-15] Their inputs and endpoints differ from this evaluation.

Comparisons should therefore preserve the conditions under which results were obtained, including supplied numerical data, full-text access and the availability of human corrections. Our case does not establish superiority over these tools or demonstrate that equivalent release controls are absent from them.

The routes without synthesis are also substantive outputs. Most included studies had no quantitative route under the frozen contracts. This should not be interpreted as absence of evidence or validation of every exclusion from synthesis. The report-level reconciliation contains categorical reasons, whereas the study-level file assigns a shared code to all 291 studies without a quantitative route, without individual narrative reasons. Future evaluation should distinguish justified non-synthesis from unnecessarily restrictive routing. Likewise, separating prognostic association, causal- intervention status and individual applicability describes the scope of evidence; it does not demonstrate that the evidence improves decisions or care. At report level, the reconciliation assigned 103 reports to incomplete source numerical data, 230 to quantitative data that could not be synthesized, 73 to direction-only synthesis and 48 to domain-level pooled synthesis. These report counts address a different unit from the study routes and must not be interchanged. Their separation is useful for examining why evidence leaves the quantitative pipeline without assuming that each routing decision was correct.

The primary limitation is that the development review was also the evaluation case. The design was retrospective and non-blinded, with no held-out domain, external comparator or independently constructed human reference. It cannot estimate comparative accuracy, non-inferiority or generalizability. Training-data contamination and prompt sensitivity were not assessed. Shared specification assumptions may affect both statistical implementations, and the 1,217 comparisons are not independent validation observations. Dependence checks rely on investigator judgments, while exposure to shared recommendations may anchor human confirmation. Responsible-use and reporting guidance support disclosing these boundaries rather than treating human approval as external validation.[22, 23, 40] No reference standard was available for estimating screening or extraction sensitivity, specificity or work saved over sampling. Source fragments that could be mechanically matched represented only part of the principal evidence. Neither agreement on recommendations nor successful compilation fills this reference-standard gap.

The event registry was reconstructed after heterogeneous audit cycles and cannot estimate failure incidence, detection sensitivity or errors prevented. Twenty-four Target 4 findings excluded by the counting rule remain in the crosswalk. Incomplete source-fragment verification, an unfinished audit, historical manifest discrepancies and eight disclosed limitations constrain the release claims. Four unsuccessful autonomous certification targets further document the extent of human intervention.

We did not isolate the contributions of LLMs, deterministic contracts and investigators through comparison or ablation. Missing runtime builds prevent exact repetition of the full agent workflow, even though frozen-output derivations and statistical engines are available.

The next evaluation should freeze the system before use in a new review, construct an independent reference under the same protocol and include a credible manual or automated comparator.

Endpoints should assess consequential errors, evidence-supported answer coverage, unjustified synthesis refusals and expert verification work. The documented defects can inform a controlled challenge set alongside unseen source material. Prospective updates, costs and clinical utility require separate studies; no workload, financial or patient benefit was measured here.

Supplementary Figure S2 distinguishes this evaluation and the companion clinical manuscript from those planned comparisons.

## Conclusions

This retrospective study examined 50 documented root-cause events in one completed systematic review; 12 had changed pooled results before correction. The four error pathways connect source interpretation and dependence judgments to analysis membership, statistical results, certainty and released files. Their implemented controls provide concrete rules for other review teams to inspect and test. Independent prospective comparisons are needed to determine whether these controls reduce consequential errors or reviewer work.

## Supporting information

Additional file 2: Reporting applicability checklist

## Data Availability

Frozen release SR-49523c19b885c87a and the supporting case-evaluation materials are archived in the paper1m directory of https://github.com/yinchuan123/social-connection-surgery-meta, version v1.3 (DOI 10.5281/zenodo.22766955; concept DOI 10.5281/zenodo.22230190). Materials include source and identity registries, analysis contracts, correction and supersession ledgers, release checks, the AER instance, figure-source data, deterministic R1 derivation code and both statistical engines. Code is licensed under Apache 2.0; data and documentation under CC BY 4.0. Copyrighted full texts, long verbatim excerpts, credentials and signed documents are excluded; restricted sources are identified bibliographically.

https://doi.org/10.5281/zenodo.22766955

## List of abbreviations

AER, accountable evidence release; AI, artificial intelligence; CI, confidence interval; GRADE, Grading of Recommendations Assessment, Development and Evaluation; HR, hazard ratio; LLM, large language model; OR, odds ratio; PI, principal investigator; PRISMA, Preferred Reporting Items for Systematic Reviews and Meta-Analyses; PROSPERO, International Prospective Register of Systematic Reviews; QUIPS, Quality In Prognosis Studies; REML, restricted maximum likelihood.

## Declarations

### Ethics approval and consent to participate

This study analysed published literature, aggregate study-level data, and deidentified system-process artifacts. No identifiable individual-level participant data were collected or analysed. Institutional ethics approval and informed consent were therefore not required.

## Consent for publication

Not applicable. This manuscript contains no identifiable individual participant information.

## Availability of data and materials

Frozen release SR-49523c19b885c87a and the supporting case-evaluation materials are archived in the paper1m directory of https://github.com/yinchuan123/social-connection-surgery-meta, version v1.3 (DOI 10.5281/zenodo.22766955; concept DOI 10.5281/zenodo.22230190). Materials include source and identity registries, analysis contracts, correction and supersession ledgers, release checks, the AER instance, figure-source data, deterministic R1 derivation code and both statistical engines. Code is licensed under Apache 2.0; data and documentation under CC BY 4.0.

Copyrighted full texts, long verbatim excerpts, credentials and signed documents are excluded; restricted sources are identified bibliographically.

## Authors’ contributions

CY: Conceptualization, Methodology, Investigation, Data curation, Formal analysis, Validation, Visualization, Writing – original draft, Writing – review & editing, Supervision, Project administration, Resources.

ZJ: Methodology, Investigation, Validation, Data curation, Writing – review & editing. His contributions included review of principal sources, source-grounded confirmation, evaluation of methodological outputs and substantive revision.

ZZ: Conceptualization, Methodology, Supervision, Validation, Writing – review & editing. His contributions included methodological and supervisory guidance, interpretation and critical revision.

## Acknowledgements

Not applicable.

## Use of artificial intelligence

Anthropic Claude Code was used from 13 July to 15 September 2026 for screening, extraction, source-based adjudication recommendations, code development, workflow coordination, audits and writing under investigator directives and locked specifications. OpenAI ChatGPT was used for earlier strategic advice; model versions and dates were not retained. From 16 to 22 September 2026, OpenAI ChatGPT with Codex tools and OpenAI Codex were used for language editing, bilingual alignment, literature comparisons, reference and declaration checks, document formatting and consistency checks. Figures 1–3 and Supplementary Figures S1–S3 were plotted from project specifications and frozen data using AI-assisted Matplotlib code; earlier versions used Graphviz and Matplotlib. Contributor percentages and point positions were calculated from existing counts. No clinical evidence extraction or statistical synthesis was rerun during this editorial work, and scientific inputs and statistical results were unchanged. On 28 September 2026, OpenAI Codex assisted with language revision, journal adaptation, reference renumbering, cross-reference checks and document formatting. The exact model snapshot and runtime build were not recorded. No scientific data were added or analyses rerun during this revision. Appendix S1 records the available model identifiers, dates, tasks and reproducibility limitations. The authors are responsible for the content and final manuscript. No AI system is an author. On 7 October 2026, OpenAI Codex assisted with revision of the methods manuscript, including the error-focused narrative, a case- summary table derived from the existing records, a reporting applicability map, cross-reference checks and formatting. The figure set was consolidated around workflow architecture, evidence- count denominators and historical failure events; the clinical selection flow is cited from the companion manuscript. No study data, independent validation experiment or statistical analysis was added. On 9 October 2026, OpenAI Codex assisted with methodological framing, relocation of event-identification methods, verification of historical correction entries, language editing, cross-reference checks and document formatting. No new source data, independent validation experiment or statistical analysis was added.

### Registration and related work

The underlying prognostic review was registered in PROSPERO (CRD420261449181). A prior version of the present manuscript was posted on medRxiv on 22 September 2026 (doi:10.64898/2026.09.21.26363538). The companion clinical manuscript uses the same frozen release to address clinical associations and certainty (doi:10.64898/2026.09.15.26363083). It was submitted to JAMA Surgery (SUR26-2789). The shared release does not provide independent replication or external validation. The clinical review’s selection flow and principal clinical effect estimates are reported in the companion manuscript; this methods paper examines processing errors, correction records and internal release checks.

## Additional files

Additional file 1. Supplementary material (PDF): Supplementary Tables S1–S11, Supplementary Figures S1–S3, Appendix S1 and supplementary notes. References use S-prefixed numbers.

Additional file 2. Reporting applicability checklist (PDF): PRISMA 2020 item-level mapping for the retrospective methodological case evaluation and its underlying clinical review.

