## Additional file 1: Supplementary material for "Evidence-processing errors and their correction in an LLM-assisted systematic review: a retrospective methodological case study"

Scientific release: SR-49523c19b885c87a. Registration: PROSPERO CRD420261449181. These supplementary materials describe the evidence compiler, its release evaluation and the methodological context.

Contents: Supplementary Tables S1–S11; Supplementary Figures S1–S3; Appendix S1 (AI systems, prompt governance and human oversight); Supplementary notes; Supplementary references. References in this supplement use S-prefixed numbers and are independent of the main manuscript bibliography.

**Supplementary Table S1. The 14 modules of the evidence compiler and the principal artifact produced by each module.**

| ID | Module | Responsibility | Primary artifact |
|---|---|---|---|
| **M01** | Protocol representation | represents the registered protocol as machine-readable eligibility, outcome and estimand contracts | PROSPERO record and frozen protocol YAML |
| **M02** | Task-conditioned progressive evidence distillation (D0–D5) | reduces records to reports, studies and estimates in stages, with each stage guided by the task the next stage must perform | distillation ledgers |
| **M03** | Isolated Reviewer A and Reviewer B | two independent extraction passes, each unable to see the other's output | dual-review ledgers |
| **M04** | Evidence-grounded adjudication | a third pass resolves disagreement against printed source text only | adjudication ledgers |
| **M05** | Source and field precedence | SOURCE_ADJUDICATED > SOURCE_VERIFIED > SOURCE_EXTRACTED > CONTROLLED_NORMALIZATION > DETECTOR_DERIVED > UNKNOWN | precedence contract |
| **M06** | Identity graph | Record to Report to Study to CohortEntity to DependenceCluster to AnalysisWeightUnit | FINAL_IDENTITY_AND_WEIGHT_RECONCILIATION.csv |
| **M07** | Evidence resolution and synthesis routing | routes each included study to one of four terminal study-level routes (pooled meta-analysis, replication target, not poolable, or mapped without a current quantitative synthesis route). Directional synthesis is recorded at the dependence-cluster level (98 of 421 clusters). | STUDY_TERMINAL_EVIDENCE_ROUTES.csv |
| **M08** | Deterministic statistical engine | fixed-effect and REML random-effects with modified Hartung–Knapp intervals, prediction intervals, leave-one-out and a multiverse over admissible preselections | AUTHORITATIVE_META_ANALYSIS_REGISTRY_v3.csv |
| **M09** | Result-level QUIPS and certainty compiler | assesses risk of bias per result, not per study, and compiles formal prognostic certainty per claim | RESULT_LEVEL_QUIPS_FINAL.csv; CLAIM_SPECIFIC_CERTAINTY_REGISTRY_FINAL.csv |
| **M10** | Correction and supersession system | records each correction as an event with a downstream rebuild status. Superseded artifacts are marked and excluded from distribution. | PHASE877R_FINAL_CORRECTION_EVENTS.csv; SUPERSESSION_MANIFEST_FINAL.csv |
| **M11** | Release compiler | one deterministic build regenerates every derived artifact and leaves frozen inputs untouched | RELEASE_MANIFEST.json |
| **M12** | Separate statistical implementation | a separately implemented engine within the same project recomputes the published quantities | INDEPENDENT_STATISTICAL_REPLICATION_FINAL.csv |
| **M13** | Root/package parity | manuscript packages are compiled from the release root and compared byte for byte | live SHA-256 comparison |
| **M14** | Bounded human confirmation and PI sign-off | two people confirm the principal source records. The principal investigator (PI) adjudicates every escalation and signs the release. | FINAL_HUMAN_CONFIRMATION_REGISTRY.csv; FINAL_SIGNED_APPROVAL_REGISTRY.csv |

**Supplementary Table S2. Automation and oversight as recorded in the signed-release governance table. The extraction row lists “third-pass adjudication from source” under “Human component” but does not name the adjudicator. The final two-person human confirmation is a separate step.**

| Stage | Automated component | Human component | Control |
|---|---|---|---|
| Screening and eligibility | system | PI spot adjudication on escalation | eligibility contract frozen before screening |
| Extraction of printed values | system (dual pass) | third-pass adjudication from source | precedence contract; no auto-fill |
| Identity and dependence clustering | system | PI directives established two cluster edges | one weight per cluster per analysis |
| Estimate preselection | system (13-level hierarchy) | PI adjudicated level semantics and ties | a level is used for selection only when every candidate can be assessed |
| Statistical estimation | system (deterministic) | none | separate statistical implementation within the same project |
| Risk of bias and certainty | system (result level) | PI adjudicated domain criteria and froze the protocol | no formal GRADE below the family level |
| Abstention decisions | system | PI reviewed each abstention | thresholds prespecified; abstention leaves a visible row |
| Escalation of ambiguous decisions | automated flagging for PI review; no automated final decision | PI decides | escalation card records both options and their measured consequences |
| Principal source confirmation | automated recommendations | two people confirm | recommendation-assisted, not blinded independent review |
| Release authorisation | automated compilation and release checks | PI signs | signature archived with SHA-256 |

**Supplementary Table S3. Analysis identities and membership checks for the five principal families. Identifiers, membership hashes and counts link each family to the frozen release for audit and reproduction. k denotes analysis weight units within an analysis. Clinical effect estimates and their interpretation are reported in the companion clinical manuscript (medRxiv doi:10.64898/2026.09.15.26363083). The corresponding frozen statistical outputs remain available in the public archive (doi:10.5281/zenodo.22766955). These shared records are not an independent validation dataset.**

| Field | Overall survival | Early mortality | Non-home discharge | Readmission | Complications |
|---|---|---|---|---|---|
| **PrincipalFamilyID** | SC-POSTOPERATIVE-OVERALL-SURVIVAL-HR-PRIMARY | JAMA-F2 | SC-NON-HOME-DISCHARGE-OR | SC-UNPLANNED-READMISSION-OR-FULL | JAMA-F4 |
| **SourceAnalysisID** | F1_SPLIT_A | JAMA-F2 | JAMA-F3 | JAMA-F6-C2 | JAMA-F4 |
| **ExactK** | 9 | 8 | 9 | 9 | 4 |
| **MembershipHash** | 8a13260f331584c3 | 28d578c0d38cc3aa | 1e9e14e771e65645 | 3275fe26473476a6 | dbc240abea1834a9 |
| **MembershipN** | 9 | 8 | 9 | 9 | 4 |
| **ReleaseBuildID** | SR-49523c19b885c87a | SR-49523c19b885c87a | SR-49523c19b885c87a | SR-49523c19b885c87a | SR-49523c19b885c87a |

**Supplementary Table S4a. Root-cause classes of the 50 documented events.**

| Root-cause class | Distinct events | % of 50 |
|---|---|---|
| AUDIT_HARNESS_FAILURE | 6 | 12.0% |
| OUTCOME_IDENTITY | 6 | 12.0% |
| COHORT_DEPENDENCE | 5 | 10.0% |
| ARTIFACT_LINEAGE | 4 | 8.0% |
| ESTIMATE_SELECTION | 4 | 8.0% |
| EVIDENCE_EXTRACTION | 4 | 8.0% |
| EXPOSURE_OR_COMPARATOR_ORIENTATION | 3 | 6.0% |
| STATISTICAL_IMPLEMENTATION | 3 | 6.0% |
| DENOMINATOR_SEMANTICS | 2 | 4.0% |
| EFFECT_SCALE_OR_TRANSFORMATION | 2 | 4.0% |
| HUMAN_GOVERNANCE | 2 | 4.0% |
| ROOT_PACKAGE_PARITY | 2 | 4.0% |
| SOURCE_IDENTITY | 2 | 4.0% |
| TIME_ORIGIN | 2 | 4.0% |
| CORRECTION_PROPAGATION | 1 | 2.0% |
| DEFAULT_AS_VERDICT | 1 | 2.0% |
| MODEL_SELECTION_TO_ROB_PROPAGATION | 1 | 2.0% |

**Supplementary Table S4b. Shared failure modes annotated across events. These are labels only; an event with a shared mode is still counted once.**

| Shared failure mode (cross-cutting label; not an additional event) | Events |
|---|---|
| SILENT_ZERO_HIT_JOIN | 3 |
| SUPERSEDED_ARTIFACT_LEAKAGE | 3 |
| AUDITED_OBJECT_MUTATED | 2 |
| COMPARATOR_ARM_DECIDES_POLARITY | 2 |
| CONSTANT_CRITERION_NEVER_DISCRIMINATES | 2 |
| GATE_THAT_CANNOT_FAIL | 2 |
| REGEX_WORD_BOUNDARY | 2 |
| DEFAULT_AS_VERDICT | 1 |

**Supplementary Table S5. Six illustrative events (four described in the main Results and two supplementary cases), classified using the frozen registry. Y/N flags: Membership, k, Pooled result, Certainty domain or grade, Denominator, Package.**

**Panel A. FE-21, FE-06 and FE-11.**

| Field | FE-21 | FE-06 | FE-11 |
|---|---|---|---|
| **Event** | FE-21 | FE-06 | FE-11 |
| **Method case** | time origin (estimand) | cohort overlap / dependence | outcome and comparator direction |
| **Root-cause class** | TIME_ORIGIN | COHORT_DEPENDENCE | EXPOSURE_OR_COMPARATOR_ ORIENTATION |
| **Trigger** | Diagnosis-origin estimates carried most of the pooled weight in a postoperative-survival family. | overlap adjudication ledger was never connected to the identity graph | The whole outcome string determined polarity, so comparator wording determined the assigned direction. The adverse override was applied before the desirable-event branch. |
| **Detection** | Target 4 audit perspective P05 | Target 1 audit finding P07-01; not fixed in cycle 1; recurred in Target 2 audit P07-01 | dual full-text source-verification passes (A/B) with third-pass review; PI patch PI-877R-FINAL-PATCH-01 |
| **Consequences (M/k/P/G/D/Pk)** | Y/Y/Y/Y/N/Y | Y/Y/Y/N/Y/Y | Y/N/Y/N/N/Y |
| **Recurred** | NO | YES | NO |
| **Preventive control** | F1 estimand split A–E; the postoperative node is primary, the all-origin node is sensitivity | adjudication_enumerator compares ledger requirement against built artifact; members read from declared fields only, never regex over prose | polarity assigned using the counted arm only; comparator clause stripped first; complement forms masked before matching; no HIGHER_IS_ WORSE fallback |
| **Status** | CLOSED_FIXED_AND_VERIFIED | CLOSED_DOWNSTREAM_REBUILD_ COMPLETE | CLOSED_DOWNSTREAM_REBUILD_ COMPLETE |
| **Source evidence** | BOUNDED_RELEASE_TARGET4_FINDING_ TRIAGE.csv#P05-01 | PHASE877R_FINAL_CORRECTION_ EVENTS.csv#CE-877R-046 | PHASE877R_FINAL_CORRECTION_ EVENTS.csv#CE-877R-024; PHASE877R_ FINAL_CORRECTION_ EVENTS.csv#CE-877R-024 |

**Supplementary Table S5 (continued). Panel B. FE-38, FE-14 and FE-35. Consequence flags: Membership, k, Pooled result, Certainty domain or grade, Denominator, Package (Y/N).**

| Field | FE-38 | FE-14 | FE-35 |
|---|---|---|---|
| **Event** | FE-38 | FE-14 | FE-35 |
| **Method case** | model selection → result-level QUIPS → certainty | controlled-vocabulary matching (word boundary) | audited object mutated during audit |
| **Root-cause class** | MODEL_SELECTION_TO_ROB_ PROPAGATION | OUTCOME_IDENTITY | AUDIT_HARNESS_FAILURE |
| **Trigger** | Results at high risk of bias accounted for more than the protocol threshold of total random-effects weight. The quality-restricted set could not be constructed. | \bhome\b failed on "Nonhome" because n and h are both word characters | the object under audit was regenerated while the audit was running |
| **Detection** | live recomputation (section 10 of the correction cycle) | self-check during the F3/F6-C1 reconciliation (section 12) | self-check (process error) |
| **Consequences (M/k/P/G/D/Pk)** | N/N/N/Y/N/Y | Y/Y/Y/N/N/Y | N/N/N/N/N/N |
| **Recurred** | NO | NO | YES |
| **Preventive control** | the tier change is recorded; criteria were not relaxed to preserve the frozen grade | aggregate non-home forms tested before the frozen implementation; no other criterion relaxed | freeze the audited set and its dossiers with a Merkle hash; generation is blocked while the freeze marker is present |
| **Status** | CLOSED_DOWNSTREAM_REBUILD_ COMPLETE | CLOSED_DOWNSTREAM_REBUILD_ COMPLETE | CLOSED_DOWNSTREAM_REBUILD_ COMPLETE |
| **Source evidence** | PHASE877R_FINAL_CORRECTION_ EVENTS.csv#CE-877R-018 | PHASE877R_FINAL_CORRECTION_ EVENTS.csv#CE-877R-021 | PHASE877R_FINAL_CORRECTION_ EVENTS.csv#CE-877R-023; PHASE877R_ FINAL_CORRECTION_ EVENTS.csv#CE-877R-040 |

Rule 1: a ledger row is a duplicate when EventID and the first 60 characters of the defect description repeat (17 rows removed; 63 rows → 46 records).

Rule 2: an EventID that carries two different defect records is split by a discriminator (CE-877R-024, CE-877R-025).

Rule 3: sibling records that share one root cause are merged into one event (6 groups: FE-11 ← CE-877R-024, CE-877R-024; FE-17 ← CE-877R-016, CE-877R-026; FE-24 ← CE-877R-036, CE-877R-038, CE-877R-033; FE-28 ← CE-877R-025, CE-877R-025; FE-33 ← CE-877R-028, CE-877R-027; FE-35 ← CE-877R-040, CE-877R-023), giving 39 events from the correction ledger.

Rule 4: Target 4 audit findings are admitted only when verified as release-blocking (10 of 34; 24 not counted).

Rule 5: the incomplete separate-agent audit A is one audit-infrastructure event. Total 39 + 10 + 1 = 50.

**Supplementary Table S6. Mapping of 63 correction-ledger rows, 34 Target 4 findings and the audit programme to the 50 counted events. The full table with deduplication keys is in the deposited materials.**

| Source ledger | Row or ID | Deduplication status | Assigned event | Counted | Reason |
|---|---|---|---|---|---|
| Correction ledger | 1 | RETAINED_RECORD | FE-03 | YES | distinct defect record |
| Correction ledger | 2 | RETAINED_RECORD | FE-01 | YES | distinct defect record |
| Correction ledger | 3 | RETAINED_RECORD | FE-04 | YES | distinct defect record |
| Correction ledger | 4 | RETAINED_RECORD | FE-29 | YES | distinct defect record |
| Correction ledger | 5 | RETAINED_RECORD | FE-20 | YES | distinct defect record |
| Correction ledger | 6 | RETAINED_RECORD | FE-15 | YES | distinct defect record |
| Correction ledger | 7 | RETAINED_RECORD | FE-22 | YES | distinct defect record |
| Correction ledger | 8 | RETAINED_RECORD | FE-23 | YES | distinct defect record |
| Correction ledger | 9 | RETAINED_RECORD | FE-27 | YES | distinct defect record |
| Correction ledger | 10 | RETAINED_RECORD | FE-30 | YES | distinct defect record |
| Correction ledger | 11 | RETAINED_RECORD | FE-31 | YES | distinct defect record |
| Correction ledger | 12 | RETAINED_RECORD | FE-02 | YES | distinct defect record |
| Correction ledger | 13 | RETAINED_RECORD | FE-05 | YES | distinct defect record |
| Correction ledger | 14 | RETAINED_RECORD | FE-07 | YES | distinct defect record |
| Correction ledger | 15 | RETAINED_RECORD | FE-16 | YES | distinct defect record |
| Correction ledger | 16 | RETAINED_RECORD | FE-17 | YES_AS_PART_OF_MERGED_EVENT | distinct defect record; merged with sibling records sharing one root cause |
| Correction ledger | 17 | RETAINED_RECORD | FE-48 | YES | distinct defect record |
| Correction ledger | 18 | RETAINED_RECORD | FE-38 | YES | distinct defect record |

| Source ledger | Row or ID | Deduplication status | Assigned event | Counted | Reason |
|---|---|---|---|---|---|
| Correction ledger | 19 | RETAINED_RECORD | FE-18 | YES | distinct defect record |
| Correction ledger | 20 | RETAINED_RECORD | FE-14 | YES | distinct defect record |
| Correction ledger | 21 | RETAINED_RECORD | FE-08 | YES | distinct defect record |
| Correction ledger | 22 | RETAINED_RECORD | FE-06 | YES | distinct defect record |
| Correction ledger | 23 | RETAINED_RECORD | FE-47 | YES | distinct defect record |
| Correction ledger | 24 | RETAINED_RECORD | FE-40 | YES | distinct defect record |
| Correction ledger | 25 | RETAINED_RECORD | FE-32 | YES | distinct defect record |
| Correction ledger | 26 | RETAINED_RECORD | FE-24 | YES_AS_PART_OF_MERGED_EVENT | distinct defect record; merged with sibling records sharing one root cause |
| Correction ledger | 27 | RETAINED_RECORD | FE-25 | YES | distinct defect record |
| Correction ledger | 28 | RETAINED_RECORD | FE-24 | YES_AS_PART_OF_MERGED_EVENT | distinct defect record; merged with sibling records sharing one root cause |
| Correction ledger | 29 | RETAINED_RECORD | FE-12 | YES | distinct defect record |
| Correction ledger | 30 | RETAINED_RECORD | FE-35 | YES_AS_PART_OF_MERGED_EVENT | distinct defect record; merged with sibling records sharing one root cause |
| Correction ledger | 31 | RETAINED_RECORD | FE-13 | YES | distinct defect record |
| Correction ledger | 32 | RETAINED_RECORD | FE-34 | YES | distinct defect record |
| Correction ledger | 33 | RETAINED_RECORD | FE-24 | YES_AS_PART_OF_MERGED_EVENT | distinct defect record; merged with sibling records sharing one root cause |
| Correction ledger | 34 | RETAINED_RECORD | FE-26 | YES | distinct defect record |
| Correction ledger | 35 | RETAINED_RECORD | FE-33 | YES_AS_PART_OF_MERGED_EVENT | distinct defect record; merged with sibling records sharing one root cause |
| Correction ledger | 36 | RETAINED_RECORD | FE-49 | YES | distinct defect record |
| Correction ledger | 37 | RETAINED_RECORD | FE-09 | YES | distinct defect record |
| Correction ledger | 38 | RETAINED_RECORD | FE-36 | YES | distinct defect record |
| Correction ledger | 39 | RETAINED_RECORD | FE-11 | YES_AS_PART_OF_MERGED_EVENT | distinct defect record; EventID carries two different defects, split by discriminator; merged with sibling records sharing one root cause |
| Correction ledger | 40 | RETAINED_RECORD | FE-28 | YES_AS_PART_OF_MERGED_EVENT | distinct defect record; EventID carries two different defects, split by discriminator; merged with sibling records sharing one root cause |

| Source ledger | Row or ID | Deduplication status | Assigned event | Counted | Reason |
|---|---|---|---|---|---|
| Correction ledger | 41 | DUPLICATE_ROW_OF_RECORD | FE-06 | NO | byte-level repeat of the same EventID and defect text; counted once |
| Correction ledger | 42 | DUPLICATE_ROW_OF_RECORD | FE-47 | NO | byte-level repeat of the same EventID and defect text; counted once |
| Correction ledger | 43 | DUPLICATE_ROW_OF_RECORD | FE-40 | NO | byte-level repeat of the same EventID and defect text; counted once |
| Correction ledger | 44 | DUPLICATE_ROW_OF_RECORD | FE-32 | NO | byte-level repeat of the same EventID and defect text; counted once |
| Correction ledger | 45 | DUPLICATE_ROW_OF_RECORD | FE-24 | NO | byte-level repeat of the same EventID and defect text; counted once |
| Correction ledger | 46 | DUPLICATE_ROW_OF_RECORD | FE-25 | NO | byte-level repeat of the same EventID and defect text; counted once |
| Correction ledger | 47 | DUPLICATE_ROW_OF_RECORD | FE-24 | NO | byte-level repeat of the same EventID and defect text; counted once |
| Correction ledger | 48 | DUPLICATE_ROW_OF_RECORD | FE-12 | NO | byte-level repeat of the same EventID and defect text; counted once |
| Correction ledger | 49 | DUPLICATE_ROW_OF_RECORD | FE-35 | NO | byte-level repeat of the same EventID and defect text; counted once |
| Correction ledger | 50 | DUPLICATE_ROW_OF_RECORD | FE-13 | NO | byte-level repeat of the same EventID and defect text; counted once |
| Correction ledger | 51 | DUPLICATE_ROW_OF_RECORD | FE-34 | NO | byte-level repeat of the same EventID and defect text; counted once |
| Correction ledger | 52 | DUPLICATE_ROW_OF_RECORD | FE-24 | NO | byte-level repeat of the same EventID and defect text; counted once |
| Correction ledger | 53 | DUPLICATE_ROW_OF_RECORD | FE-26 | NO | byte-level repeat of the same EventID and defect text; counted once |
| Correction ledger | 54 | DUPLICATE_ROW_OF_RECORD | FE-33 | NO | byte-level repeat of the same EventID and defect text; counted once |
| Correction ledger | 55 | DUPLICATE_ROW_OF_RECORD | FE-49 | NO | byte-level repeat of the same EventID and defect text; counted once |
| Correction ledger | 56 | DUPLICATE_ROW_OF_RECORD | FE-09 | NO | byte-level repeat of the same EventID and defect text; counted once |

| Source ledger | Row or ID | Deduplication status | Assigned event | Counted | Reason |
|---|---|---|---|---|---|
| Correction ledger | 57 | DUPLICATE_ROW_OF_RECORD | FE-36 | NO | byte-level repeat of the same EventID and defect text; counted once |
| Correction ledger | 58 | RETAINED_RECORD | FE-11 | YES_AS_PART_OF_MERGED_EVENT | distinct defect record; EventID carries two different defects, split by discriminator; merged with sibling records sharing one root cause |
| Correction ledger | 59 | RETAINED_RECORD | FE-28 | YES_AS_PART_OF_MERGED_EVENT | distinct defect record; EventID carries two different defects, split by discriminator; merged with sibling records sharing one root cause |
| Correction ledger | 60 | RETAINED_RECORD | FE-17 | YES_AS_PART_OF_MERGED_EVENT | distinct defect record; merged with sibling records sharing one root cause |
| Correction ledger | 61 | RETAINED_RECORD | FE-33 | YES_AS_PART_OF_MERGED_EVENT | distinct defect record; merged with sibling records sharing one root cause |
| Correction ledger | 62 | RETAINED_RECORD | FE-35 | YES_AS_PART_OF_MERGED_EVENT | distinct defect record; merged with sibling records sharing one root cause |
| Correction ledger | 63 | RETAINED_RECORD | FE-46 | YES | distinct defect record |
| Target 4 triage | P01-02 | TARGET4_FINDING | FE-41 | YES | verified release-blocking Target 4 finding (VerifiedReleaseBlocking=True) admitted as an additional root-cause event |
| Target 4 triage | P01-04 | TARGET4_FINDING | – | NO | not counted: VerifiedReleaseBlocking=False; triage=MAIN_RELEASE_CRITICAL; disposition=MUST_FIX_AND_VERIFY — audit allegation handled inside the bounded release but not admitted as a distinct terminally-confirmed root cause under the frozen counting rule |
| Target 4 triage | P01-05 | TARGET4_FINDING | – | NO | not counted: VerifiedReleaseBlocking=False; triage=MAIN_RELEASE_CRITICAL; disposition=MUST_FIX_AND_VERIFY — audit allegation handled inside the bounded release but not admitted as a distinct terminally-confirmed root cause under the frozen counting rule |
| Target 4 triage | P01-07 | TARGET4_FINDING | – | NO | not counted: VerifiedReleaseBlocking=False; triage=MAIN_RELEASE_CRITICAL; disposition=MUST_FIX_AND_VERIFY — audit allegation handled inside the bounded release but not admitted as a distinct terminally-confirmed root cause under the frozen counting rule |

| Source ledger | Row or ID | Deduplication status | Assigned event | Counted | Reason |
|---|---|---|---|---|---|
| Target 4 triage | P02-01 | TARGET4_FINDING | – | NO | not counted: VerifiedReleaseBlocking=False; triage=MAIN_RELEASE_CRITICAL; disposition=MUST_FIX_AND_VERIFY — audit allegation handled inside the bounded release but not admitted as a distinct terminally-confirmed root cause under the frozen counting rule |
| Target 4 triage | P02-06 | TARGET4_FINDING | – | NO | not counted: VerifiedReleaseBlocking=False; triage=MAIN_RELEASE_CRITICAL; disposition=MUST_FIX_AND_VERIFY — audit allegation handled inside the bounded release but not admitted as a distinct terminally-confirmed root cause under the frozen counting rule |
| Target 4 triage | P02-07 | TARGET4_FINDING | – | NO | not counted: VerifiedReleaseBlocking=False; triage=MAIN_RELEASE_CRITICAL; disposition=MUST_FIX_AND_VERIFY — audit allegation handled inside the bounded release but not admitted as a distinct terminally-confirmed root cause under the frozen counting rule |
| Target 4 triage | P03-02 | TARGET4_FINDING | FE-19 | YES | verified release-blocking Target 4 finding (VerifiedReleaseBlocking=True) admitted as an additional root-cause event |
| Target 4 triage | P03-04 | TARGET4_FINDING | – | NO | not counted: VerifiedReleaseBlocking=False; triage=MAIN_RELEASE_CRITICAL; disposition=MUST_FIX_AND_VERIFY — audit allegation handled inside the bounded release but not admitted as a distinct terminally-confirmed root cause under the frozen counting rule |
| Target 4 triage | P03-05 | TARGET4_FINDING | – | NO | not counted: VerifiedReleaseBlocking=False; triage=MAIN_RELEASE_CRITICAL; disposition=MUST_FIX_AND_VERIFY — audit allegation handled inside the bounded release but not admitted as a distinct terminally-confirmed root cause under the frozen counting rule |
| Target 4 triage | P03-06 | TARGET4_FINDING | FE-43 | YES | verified release-blocking Target 4 finding (VerifiedReleaseBlocking=True) admitted as an additional root-cause event |
| Target 4 triage | P03-08 | TARGET4_FINDING | – | NO | not counted: VerifiedReleaseBlocking=False; triage=MAIN_RELEASE_CRITICAL; disposition=MUST_FIX_AND_VERIFY — audit allegation handled inside the bounded release but not admitted as a distinct terminally-confirmed root cause under the frozen counting rule |

| Source ledger | Row or ID | Deduplication status | Assigned event | Counted | Reason |
|---|---|---|---|---|---|
| Target 4 triage | P05-01 | TARGET4_FINDING | FE-21 | YES | verified release-blocking Target 4 finding (VerifiedReleaseBlocking=True) admitted as an additional root-cause event |
| Target 4 triage | P05-02 | TARGET4_FINDING | FE-50 | YES | verified release-blocking Target 4 finding (VerifiedReleaseBlocking=True) admitted as an additional root-cause event |
| Target 4 triage | P05-04 | TARGET4_FINDING | – | NO | not counted: VerifiedReleaseBlocking=False; triage=MAIN_RELEASE_CRITICAL; disposition=MUST_FIX_AND_VERIFY — audit allegation handled inside the bounded release but not admitted as a distinct terminally-confirmed root cause under the frozen counting rule |
| Target 4 triage | P05-07 | TARGET4_FINDING | – | NO | not counted: VerifiedReleaseBlocking=False; triage=MAIN_RELEASE_CRITICAL; disposition=MUST_FIX_AND_VERIFY — audit allegation handled inside the bounded release but not admitted as a distinct terminally-confirmed root cause under the frozen counting rule |
| Target 4 triage | P06-01 | TARGET4_FINDING | – | NO | not counted: VerifiedReleaseBlocking=False; triage=MAIN_RELEASE_CRITICAL; disposition=MUST_FIX_AND_VERIFY — audit allegation handled inside the bounded release but not admitted as a distinct terminally-confirmed root cause under the frozen counting rule |
| Target 4 triage | P06-02 | TARGET4_FINDING | – | NO | not counted: VerifiedReleaseBlocking=False; triage=MAIN_RELEASE_CRITICAL; disposition=MUST_FIX_AND_VERIFY — audit allegation handled inside the bounded release but not admitted as a distinct terminally-confirmed root cause under the frozen counting rule |
| Target 4 triage | P06-05 | TARGET4_FINDING | – | NO | not counted: VerifiedReleaseBlocking=False; triage=MAIN_RELEASE_CRITICAL; disposition=MUST_FIX_AND_VERIFY — audit allegation handled inside the bounded release but not admitted as a distinct terminally-confirmed root cause under the frozen counting rule |
| Target 4 triage | P06-06 | TARGET4_FINDING | – | NO | not counted: VerifiedReleaseBlocking=False; triage=MAIN_RELEASE_CRITICAL; disposition=MUST_FIX_AND_VERIFY — audit allegation handled inside the bounded release but not admitted as a distinct terminally-confirmed root cause under the frozen counting rule |

| Source ledger | Row or ID | Deduplication status | Assigned event | Counted | Reason |
|---|---|---|---|---|---|
| Target 4 triage | P07-01 | TARGET4_FINDING | – | NO | not counted: VerifiedReleaseBlocking=False; triage=MAIN_RELEASE_CRITICAL; disposition=MUST_FIX_AND_VERIFY — audit allegation handled inside the bounded release but not admitted as a distinct terminally-confirmed root cause under the frozen counting rule |
| Target 4 triage | P07-02 | TARGET4_FINDING | FE-44 | YES | verified release-blocking Target 4 finding (VerifiedReleaseBlocking=True) admitted as an additional root-cause event |
| Target 4 triage | P07-03 | TARGET4_FINDING | – | NO | not counted: VerifiedReleaseBlocking=False; triage=MAIN_RELEASE_CRITICAL; disposition=MUST_FIX_AND_VERIFY — audit allegation handled inside the bounded release but not admitted as a distinct terminally-confirmed root cause under the frozen counting rule |
| Target 4 triage | P07-04 | TARGET4_FINDING | – | NO | not counted: VerifiedReleaseBlocking=False; triage=MAIN_RELEASE_CRITICAL; disposition=MUST_FIX_AND_VERIFY — audit allegation handled inside the bounded release but not admitted as a distinct terminally-confirmed root cause under the frozen counting rule |
| Target 4 triage | P07-05 | TARGET4_FINDING | – | NO | not counted: VerifiedReleaseBlocking=False; triage=MAIN_RELEASE_CRITICAL; disposition=MUST_FIX_AND_VERIFY — audit allegation handled inside the bounded release but not admitted as a distinct terminally-confirmed root cause under the frozen counting rule |
| Target 4 triage | P07-07 | TARGET4_FINDING | – | NO | not counted: VerifiedReleaseBlocking=False; triage=MAIN_RELEASE_CRITICAL; disposition=MUST_FIX_AND_VERIFY — audit allegation handled inside the bounded release but not admitted as a distinct terminally-confirmed root cause under the frozen counting rule |
| Target 4 triage | P01-03 | TARGET4_FINDING | FE-42 | YES | verified release-blocking Target 4 finding (VerifiedReleaseBlocking=True) admitted as an additional root-cause event |
| Target 4 triage | P02-02 | TARGET4_FINDING | – | NO | not counted: VerifiedReleaseBlocking=False; triage=DIAGNOSTIC_ONLY; disposition=RECORD_IN_LIMITATION_LEDGER — audit allegation handled inside the bounded release but not admitted as a distinct terminally-confirmed root cause under the frozen counting rule |

| Source ledger | Row or ID | Deduplication status | Assigned event | Counted | Reason |
|---|---|---|---|---|---|
| Target 4 triage | P02-03 | TARGET4_FINDING | FE-39 | YES | verified release-blocking Target 4 finding (VerifiedReleaseBlocking=True) admitted as an additional root-cause event |
| Target 4 triage | P03-03 | TARGET4_FINDING | – | NO | not counted: VerifiedReleaseBlocking=False; triage=SUPPORTING_ARTIFACT_NONCRITICAL; disposition=RECORD_IN_LIMITATION_LEDGER — audit allegation handled inside the bounded release but not admitted as a distinct terminally-confirmed root cause under the frozen counting rule |
| Target 4 triage | P03-07 | TARGET4_FINDING | – | NO | not counted: VerifiedReleaseBlocking=False; triage=SUPPORTING_ARTIFACT_NONCRITICAL; disposition=RECORD_IN_LIMITATION_LEDGER — audit allegation handled inside the bounded release but not admitted as a distinct terminally-confirmed root cause under the frozen counting rule |
| Target 4 triage | P05-05 | TARGET4_FINDING | – | NO | not counted: VerifiedReleaseBlocking=False; triage=DIAGNOSTIC_ONLY; disposition=RECORD_IN_LIMITATION_LEDGER — audit allegation handled inside the bounded release but not admitted as a distinct terminally-confirmed root cause under the frozen counting rule |
| Target 4 triage | P11-A1 | TARGET4_FINDING | FE-45 | YES | verified release-blocking Target 4 finding (VerifiedReleaseBlocking=True) admitted as an additional root-cause event |
| Target 4 triage | P11-A2 | TARGET4_FINDING | FE-10 | YES | verified release-blocking Target 4 finding (VerifiedReleaseBlocking=True) admitted as an additional root-cause event |
| Audit programme | AUDIT-A | AUDIT_INFRASTRUCTURE_EVENT | FE-37 | YES | separate-agent audit A did not complete (three infrastructure failures); substituted by deterministic local checks; counted as one audit-harness event |

**Supplementary Table S7. Mapping of the 8 disclosed non-critical limitations of the release to the 4 disclosed failure events. These are different denominators and must not be added.**

| Limitation | Category | Artifact | Counted as failure event | Event | Root-cause class |
|---|---|---|---|---|---|
| P01-03 | SUPPORTING_ARTIFACT_NONCRITICAL | SUPERSESSION_MANIFEST_FINAL_v2.csv | YES | FE-42 | ARTIFACT_LINEAGE |
| P02-02 | DIAGNOSTIC_ONLY | FINAL_MODEL_SELECTION_TERMINAL_STATE_AUDIT.csv | NO | – | – |
| P02-03 | DIAGNOSTIC_ONLY | ATTRIBUTE_MISMATCH_TO_FINAL_VALUE_AUDIT.csv | YES | FE-39 | CORRECTION_PROPAGATION |
| P03-03 | SUPPORTING_ARTIFACT_NONCRITICAL | AUTHORITATIVE_META_MEMBERSHIP_v3.csv | NO | – | – |
| P03-07 | SUPPORTING_ARTIFACT_NONCRITICAL | AUTHORITATIVE_META_MEMBERSHIP_v3.csv | NO | – | – |
| P05-05 | DIAGNOSTIC_ONLY | PHASE877R_TARGET4_READONLY/PHASE877R_FINAL_CORRECTION_EVENTS.csv | NO | – | – |
| P11-A1 | SUPPORTING_ARTIFACT_NONCRITICAL | JAMA_SURGERY_RESULTS_PACKAGE_TARGET4_CANDIDATE/tables/TABLE9_directional_synthesis.csv | YES | FE-45 | ROOT_PACKAGE_PARITY |
| P11-A2 | SUPPORTING_ARTIFACT_NONCRITICAL | FINAL_DIRECTIONAL_SYNTHESIS_v2.csv | YES | FE-10 | EVIDENCE_EXTRACTION |

DIAGNOSTIC_ONLY = affects an audit/diagnostic artifact only; SUPPORTING_ARTIFACT_NONCRITICAL = supporting evidence layer, not release-blocking unless contradicting a principal-family direction.

**Supplementary Table S8. Crosswalk to the published guidance for manuscripts testing generative AI for systematic review and meta-analysis (Farotimi et al, 2026) [S1]: requirement, applicability, evidence, gap and disposition.**

| ID | Requirement | Applicability | Evidence in this study | Gap | Disposition |
|---|---|---|---|---|---|
| AIG-01 | Clear research design allowing replication and validation | APPLICABLE | design stated: retrospective, non-blinded, single case; R1 regeneration completed | no held-out domain | DESIGN_LIMITATION_DISCLOSED |
| AIG-02 | Benchmark against traditional manual methods and/or existing automated approaches | APPLICABLE | NOT_EVALUATED: no comparator arm; no performance comparison with other systems | comparator absent | NOT_EVALUATED_DESIGN_LIMITATION_DISCLOSED |
| AIG-03 | External validation on sample set(s) independent from training/development data | APPLICABLE | NOT_EVALUATED: development and evaluation on the same review | external validation absent | NOT_EVALUATED_DESIGN_LIMITATION_DISCLOSED |
| AIG-04 | Disclose training/test cross-contamination risk and estimate performance on data unlikely in training | APPLICABLE | NOT_EVALUATED: risk stated in Limitations and Appendix S1 (appraised reports are published articles that may be in the training data of the commercial models); no performance estimate on data unlikely to be in training | no estimate made | DISCLOSED_NOT_EVALUATED |
| AIG-05 | Model name/version, API versions, access dates | APPLICABLE | Claude model identifiers and dates from the execution logs, including subagent and workflow transcripts (Appendix S1) | The Claude Code runtime build identifier and earlier ChatGPT model versions and dates of use were not recorded. Manuscript-preparation use on 16–22 and 28 September 2026 is described in Appendix S1, section H. | PARTIALLY_MET_WITH_DISCLOSED_GAP |
| AIG-06 | Prompt development/testing and prompt sensitivity | APPLICABLE | specifications described in Appendix S1 (eligibility criteria, certainty protocol, human-confirmation methods and release manifest public; synthesis contracts, vocabularies and schemas deposited in v1.3; other specifications, the numbered directives and the prompt templates available from the author, not deposited); no prompt-sensitivity experiment | prompt sensitivity NOT_EVALUATED; directives and prompt templates not deposited | PARTIALLY_MET |
| AIG-07 | Temperature and configuration settings | APPLICABLE | NOT_RECORDED (agent runtime does not expose sampling settings) | not recorded | NOT_RECORDED_DISCLOSED |

| ID | Requirement | Applicability | Evidence in this study | Gap | Disposition |
|---|---|---|---|---|---|
| AIG-08 | Random seeds for data splitting/initialisation/sampling | PARTLY APPLICABLE | deterministic statistical engine (no seeds; no train/test split); language-model sampling seeds not controllable in the agent runtime (NOT_RECORDED); selection rule and seed of the risk-weighted full-text verification sample NOT_RECORDED in the inputs of this study | seeds of stochastic agent steps not recorded | PARTIALLY_MET_WITH_DISCLOSED_GAP |
| AIG-09 | Established SRMA performance metrics (WSS, NNR, precision, sensitivity, specificity) | APPLICABLE | NOT_EVALUATED: no reference standard for screening/extraction accuracy; release-integrity domains reported instead | task accuracy not measured | NOT_EVALUATED_DISCLOSED |
| AIG-10 | Human oversight method, extent of review, error handling | APPLICABLE | PI directives, escalation cards, two-person confirmation of 39 records, release sign-off, correction ledger | confirmation assisted by recommendations | MET |
| AIG-11 | Complete executable code; public datasets in established repositories | APPLICABLE | frozen release files, R1 build code, R2 engines and the other materials of this study deposited as v1.3 of the public repository (concept DOI 10.5281/zenodo.22230190) | full texts excluded (copyright); R3 not reproducible | PARTIALLY_MET |
| AIG-12 | Software versions, dependencies, hardware | APPLICABLE | The manuscript build environment is recorded in the deposited materials (v1.3, ENVIRONMENT.json), with package versions read at build time. | software environment of the release build (production and clean-room engine runs) NOT_RECORDED; hardware not material | PARTIALLY_MET |
| AIG-13 | Limitations, uncertainties, biases; unsubstantiated capability claims avoided | APPLICABLE | The manuscript states design and validation limitations. Claims were screened for unsupported assertions during manuscript preparation. | none | MET |
| AIG-14 | Conflicts of interest incl. commercial relationships | APPLICABLE | See the competing interests statement in the main manuscript. | Declarations are author-reported and were not independently verified. | AUTHOR_DECLARED |
| AIG-15 | Performance across conditions (prompts, contexts, data types) | APPLICABLE | NOT_EVALUATED: one review, one data type, one prompt set | no cross-condition evaluation | NOT_EVALUATED_DISCLOSED |

**Supplementary Table S9. Reported evaluation scopes of related systems, methods and guidance.**

| Work and source | Publication type and assessed version | Evaluation scope, inputs and human role | Relation to this study and limits |
|---|---|---|---|
| **TrialMind [S2]** | Peer-reviewed; npj Digital Medicine, 2025 | Search, screening, extraction and synthesis benchmarked on 100 oncology reviews (2,220 studies), using annotated references and human/LLM comparators. A separate human–AI workload pilot involved two participants. | The benchmark and workload pilot have different denominators. Time savings in that pilot cannot estimate this project's savings. Release-level conformance of the kind evaluated here was not reported. |
| **Manalyzer [S3]** | PREPRINT (arXiv v2 2026-01-21; no journal version found) | multi-agent end-to-end meta-analysis; benchmark of 729 papers in three non-clinical domains | screening/extraction vs expert labels; no clinical domain; release-level evaluation not reported |
| **AutoSynthesis [S4]** | Preprint; arXiv v1, 16 July 2026 | Agentic workflow from question to report, illustrated by a limited set of predominantly non-clinical reference examples, including human–AI interaction and persuasion. Pooled outputs are compared with human-produced syntheses. | The examples establish workflow feasibility under the tested conditions. They do not establish broad clinical generalizability, and independent external clinical validation was not reported. |
| **otto-SR [S5]** | Preprint; assessed full text: v3, 18 February 2026 | Screening, extraction and risk-of-bias assessment; reproduction and updating of predefined primary analyses from 12 Cochrane reviews, comparing original, automated and human-corrected synthesis outputs. | Examines downstream changes after correction and updating. The present study focuses on typed evidence identity, declared dependence, correction propagation and a frozen publication package. Comparative superiority is untested. |
| **Chen et al [S6]** | Peer-reviewed; Nature Medicine, 2026 | Clinical LLM review of 4,609 studies, classified primarily from titles and abstracts. GPT-5 screening was checked against a tie-broken human reference in 500 records. Tiering used a separate 250-record sample. | The corpus is not 4,609 expert-adjudicated full texts. Extraction was not separately validated; task validation does not establish complete numerical synthesis or release conformance. |
| **Li et al [S7]** | Peer-reviewed; RSM 17:671–692, 2026 | Extraction from 58 papers across three domains with three models and multiple prompts. Evaluation used source-derived references and an LLM, with human checking of a sample of evaluation labels. Omissions were a major error category. | Completeness and evaluator agreement have distinct denominators. Suggested shares of human effort and target precision/recall do not measure labour savings. Downstream clinical conclusions and release conformance were not evaluated. |
| **EviSearch [S8]** | Preprint; arXiv v2, 21 April 2026 | Structured clinical extraction evaluated on 667 fields against clinical annotations using an LLM evaluator. Dual extraction, page-level disagreement checks, cell attribution and a human audit interface are described; API/token use is reported. | Agreement between extractors can leave shared omissions undetected (our inference). Token use was higher than the native-PDF baseline and lower than the parsed-text baselines. Statistical dependence, clinical benefit and full-release conformance were not evaluated. |
| **INSPECT-AI / RIPE [S9]** | Preprint; assessed arXiv v1, 7 August 2026 | Provenance knowledge graphs help create and manage research-integrity assessments. Evaluation covers 95 trial reports, 140 assessments and four automated question types, compared with human assessments. | Agreement across assessment–question pairs is not fraud-detection accuracy. Human review remains necessary. Study trustworthiness, file identity/full-text validity and risk of bias are distinct objects of evaluation. |
| **Paper2Agent [S10]** | Peer-reviewed; Nature, online 16 September 2026 | Papers, code and data become executable MCP tools, with automated tests against expected execution and reference outputs. Evaluation includes 100 computational-biology papers and additional cross-domain, refusal and repository-drift tests. | Successful paper conversion and passing tool tests have different denominators. Execution and output reproduction do not establish all scientific interpretations or clinical benefit. The task depends on access to usable code and data. |

| Work and source | Publication type and assessed version | Evaluation scope, inputs and human role | Relation to this study and limits |
|---|---|---|---|
| **ARISMA [S11]** | Assessed preprint; arXiv v1, 25 August 2026; PDF states AGENTICS 2026 acceptance | Guidance for AI-assisted systematic, scoping and mapping reviews, including human oversight, logs, version tracking and reversible decisions. It proposes reporting and governance practices rather than a comparative performance evaluation. | These principles are relevant precedent for governance. ARISMA is not an endorsed PRISMA extension or evidence that an implementation meets its recommendations. The present AER instance is also not a certification standard. |
| **Khraisha et al [S12]** | PEER_REVIEWED | GPT-4 screening/extraction in one review; modest kappa; caution urged | single review; human reference partly single-reviewer |
| **Purewal et al [S13]** | Published task comparison | Screening in one chronic-pain review and pooling of five supplied datasets. The meta-analysis step received study-level mean differences, confidence intervals and weights. | Near agreement when pooling supplied values coexisted with lower screening performance. This evaluates numerical reproduction under given inputs, not the accuracy of a complete autonomous review. |
| **Sollini et al [S14]** | Published multitask comparison | Retrieval, extraction and drafting for one review under deliberately simple prompts. Tasks were evaluated separately, with human-derived inputs supplied to later stages. Two rounds compared model/service versions. | Findings depend on the tested prompting and input conditions. The two rounds are not a living-review update series or a controlled test of a fixed model version. |
| **Kim et al [S15]** | Published proof-of-concept study | Four customized GPT-4 workflows for extraction and risk-of-bias assessment, using supplied trial PDFs, with source checking, repeated runs and code-based synthesis across four selected reviews. | Human oversight, modular processing and code transparency are established components. Reported differences in heterogeneity and risk-of-bias judgments distinguish agreement from validated source and synthesis decisions. |
| **Pratte et al [S16]** | Published meta-research study | Rapid approximation of 23 included critical-care meta-analyses. 12 attempts were excluded after persistent reliance on review-level evidence. Magnitude agreement allowed a point estimate within the published 95% confidence interval, and GRADE could differ by one level. | Approximate concordance is not exact reproduction, verified study membership or clinical validity. Model processing time is not the total work required for an acceptable evidence release. |
| **Zou et al [S17]** | Published two-topic evaluation | Title-and-abstract screening, retrieval-augmented drafting and PRISMA reporting assessment in hepatology. Full-text eligibility remained manual; drafting and automated assessment used the o1 model family. | Good screening performance coexisted with numerical extraction/synthesis errors and unsupported findings. Reporting scores do not verify numerical correctness. Longitudinal release evaluation was not reported. |
| **ASReview [S18, S19]** | PEER_REVIEWED (software); v3.0.8 2026 | active-learning screening; no LLM screening documented | screening only |
| **Rayyan [S20, S21]** | SOFTWARE_PAPER + independent evaluation of ratings | ML ranking; LLM features (ResearchPilot) VENDOR-DESCRIBED, no peer-reviewed evaluation found | NOT_PUBLICLY_DOCUMENTED for LLM features |
| **DistillerSR [S22, S23]** | PEER_REVIEWED evaluations of screening features | AI rerank/classifiers; GenAI extraction (SEE) VENDOR-DESCRIBED | GenAI extraction: NOT_PUBLICLY_DOCUMENTED |

| Work and source | Publication type and assessed version | Evaluation scope, inputs and human role | Relation to this study and limits |
|---|---|---|---|
| **Elicit [S24, S25]** | PEER_REVIEWED evaluations (search/screening; extraction feasibility) | vendor systematic-review product with internal Cochrane-based evaluation (VENDOR-DESCRIBED) | no peer-reviewed end-to-end evaluation found |
| **Nested Knowledge [S26]** | PEER_REVIEWED validation (vendor-affiliated author) | Robot Screener / Smart Screener / extraction; living review features VENDOR-DESCRIBED | independence of evaluation limited |
| **Cochrane randomized controlled trial classifier [S27]** | PEER_REVIEWED classifier evaluation | Machine-learning randomized controlled trial classification evaluated for Cochrane Reviews (Thomas et al, 2021). | Evaluation of a screening classifier; it does not establish the performance of the full EPPI-Reviewer platform. |
| **Reason et al [S28]** | Published adjacent-domain feasibility study; not SR validation | GPT-4 construction of two prespecified health-economic models, with repeated generation, human technical checking and comparison with published base-case outputs. | Provides a precedent for using an LLM to generate structured model code. The treatment cost/QALY ratios do not measure the cost-effectiveness of the LLM workflow or the present evidence compiler. |
| **Compiling prompts [S29]** | PEER_REVIEWED (MethodsX 2026) following arXiv:2509.00038 | prompt compilation/optimisation protocol for LLM review workflows | screening only; single dataset |
| **RAISE [S30, S31]** | ORGANISATIONAL_DOCUMENT (living draft v3, 2026-03-13) + peer-reviewed position statement | responsible-use recommendations; human responsibility; transparent reporting | Living draft: not peer-reviewed; evolving. |
| **Journal guidance (Farotimi et al) [S1]** | GUIDANCE (editorial) | requirements for manuscripts testing generative AI | this study does not meet comparator/external-validation expectations |
| **TRIPOD-LLM [S32]** | PEER_REVIEWED reporting guideline (EQUATOR-listed) | reporting of LLM studies | partial applicability to system evaluation |
| **PRISMA-trAIce [S33, S34]** | PEER_REVIEWED proposal; NOT an endorsed PRISMA extension (PRISMA Executive letter 2026) | proposed checklist for AI use in SLRs | not endorsed; not EQUATOR-registered |

| Work and source | Publication type and assessed version | Evaluation scope, inputs and human role | Relation to this study and limits |
|---|---|---|---|
| **Yao and Yu [S35]** | Perspective; Article in Press, 19 September 2026 | Proposes linked claim, evidence, link and audit records, with the answer, evidence package and workflow profile as the evaluation unit. Covers evidence adequacy, distinct evidence bases, source versions, unresolved states, package reproducibility, change control and human review. | Conceptual framework with a schematic example. Shares source, version and review records with AER; prospective work-system validation remains required. Meta-analysis-specific weight invariants and event-linked pooled-result changes are NOT_REPORTED_IN_CHECKED_SOURCE. The present case links these statistical objects to correction and release records; no comparative performance was tested. |
| **MRER; Gao et al [S36]** | Article in Press, 19 September 2026; main article checked; supplement and code not checked | Medical multiple-choice QA on three benchmarks using textbook retrieval, primarily with Llama-3.1-8B-Instruct. Reports accuracy comparisons, ablations, time/token use and blinded expert ratings of 60 cases. | PICO-guided retrieval, accumulated evidence and auditor feedback are precedents. Evaluation concerns answers and reasoning trajectories. The present case examines statistical contributions, correction events and a fixed synthesis release. Synthesis-release conformance was not an evaluated endpoint in the checked main article; unexamined implementation features remain NOT_VERIFIED. No performance comparison was made. |

Existing sources were assessed through 17 September 2026; the Yao and Yu and MRER main articles were additionally assessed in full on 20 September 2026. Entries describe the assessed reports, their evaluation conditions, human involvement and limitations; they do not catalogue every system capability. The otto-SR full-text assessment used v3. Version v4 (4 May 2026) was checked only at the metadata and abstract level. VENDOR-DESCRIBED identifies vendor documentation. NOT_PUBLICLY_DOCUMENTED means that no public evaluation was located in the recorded assessment. NOT_REPORTED_IN_CHECKED_SOURCE and NOT_VERIFIED identify limits of the source assessment, not absent capabilities. No head-to-head comparison with the present system was performed. S-prefixed reference numbers refer to the bibliography of this supplement, independently of the main manuscript.

**Supplementary Table S10. Clinical translation considerations and the scope of the present evidence-release framework.**

| Clinical translation consideration | Corresponding artifacts in this study | Evidence in this study and its limits | Evaluation still required |
|---|---|---|---|
| **Preserve the clinical question and the meaning of evidence** | Typed source and estimate records; outcome/time-origin fields; the dependence graph; analysis contracts and correction ledgers. | Implemented in one prognostic review. Recorded failures show why these links matter. Graph checks cannot establish that every source judgment or dependence relation is correct. | Independent assessment of consequential errors in inference and of coverage across other questions, populations and source formats. |
| **Keep evidence current and assess stability over time** | A frozen release identifier, correction history, supersession records and explicit analysis states. | The study evaluated one fixed release. It did not prospectively add new studies or measure the time needed for repeated updates, evidence stability over time or resources used for updates. | Longitudinal observation of incoming evidence, corrections, changes in conclusions and the work needed to produce acceptable updates. |
| **Establish applicability to patients and intended populations** | Separate evidence levels for association, magnitude, transportability, individual applicability and causal-intervention status. | These categories constrain interpretation but are not validated patient-specific recommendations. The study did not recruit patients or evaluate representativeness, comprehension or outcomes. | Prospective evaluation of applicability, selection effects, patient understanding, shared decision-making and clinical benefit in the intended care setting. |
| **Retain meaningful human responsibility** | PI adjudication and release authorization; recommendation-assisted two-person confirmation of principal records. | Human roles are documented within evidence production. Confirmation was not a blinded independent reference, and the study did not test clinical oversight or accountability arrangements. | Evaluation of supervision, escalation, handoff, responsibility and the effects of clinician and patient interaction during actual use. |
| **Control versions and identify what can be reproduced** | Versioned evidence artifacts, deterministic derivation inputs/code, two statistical implementations and publication-package parity checks. | Release identity and numerical reproduction within the stated scope were checked. Missing runtime identifiers and nondeterministic agent outputs limit strict R3 reproduction. The release does not freeze an entire clinical AI service. | Stable model and deployment definitions during evaluation, documented changes, and monitoring after updates to evidence or software. |
| **Integrate with clinical workflows and monitor safety** | Inspectable provenance, explicit non-synthesis states and documented correction consequences. | These artifacts could inform downstream systems. The study did not evaluate EHR integration, clinical workflow, patient trust, harm detection or safety monitoring. | Prospective assessment of workflow integration, handoffs, usability, safety and patient outcomes under real operating conditions. |
| **Account for resources and retain necessary scientific records** | AI execution summaries and retained source, audit, correction and supersession artifacts. | The study did not collect a complete account of workflow costs or a comparable manual or automated baseline. Resource savings and cost-effectiveness were not established. | Measurement of expert preparation, adjudication, correction and checking time; computation and maintenance; and storage/retention costs, alongside quality and coverage. |

This table relates existing study artifacts to the clinical translation considerations discussed by Schaekermann et al [S37] and to the living-evidence and resource issues described by Cao and Moher [S38], Wall et al [S39] and Reason et al [S28]. It adds no new experiment, validated deployment standard or prespecified study protocol. Checks on evidence production do not establish the effectiveness or safety of a clinical AI intervention.

**Supplementary Table S11. Implemented components and limits.**

| Component | Problem | Existing approaches | System implementation | Release evidence | Validation gap |
|---|---|---|---|---|---|
| **1. Evaluation of a complete, versioned release** | Task metrics alone may miss defects that span stages of the released package | Task benchmarks; review updates; executable artifacts; answer–evidence-package–workflow evaluation | Frozen release with a manifest, release gates, hashed inputs and explicit status terms | 9/9 gates; 50 root-cause events (46 closed, 4 disclosed); 12 had changed a pooled result before correction | No external reference for release correctness. One retrospective, non-blinded case. |
| **2. Typed evidence identity and dependence** | Study counts depend on the unit. Overlapping registries may receive duplicate weight, and weight units vary by analysis. | Report and cohort linkage; provenance graphs; distinct evidence bases; reviewer judgment | Seven linked identifier types; one weight per dependence cluster per analysis; selected-estimate count = weight-unit count = k; source-precedence states | 454 reports / 445 studies / 441 cohort entities / 421 clusters / 216 weight units / 113 estimates; D2 5/5; D3 5/5 | The graph is adjudicated, not measured. The invariant holds by construction. Source review and PI adjudication found duplicate weights; invariant tests did not. |
| **3. Correction propagation and artifact supersession** | Corrections may leave stale outputs; superseded tables may be distributed | Versioned logs; correction checks; living updates; reversible governance; source-update management and change control | Correction events link consequences and rebuild status; manifest tracks artifact status; deterministic build and root/package byte parity | 18 superseded + 2 invalidated of 51 artifacts; none shipped under the final manifest. A shipped table was marked superseded in an earlier manifest (FE-45). D5 94/94; content checks 83/83 | Parity checks cover only bytes and declared status. Unrecorded corrections cannot be checked. Cross-platform determinism is not claimed. |

The release status vocabulary prohibits the label “fully autonomous clean release”. k denotes the number of analysis weight units within an analysis.

### Supplementary Figure S1

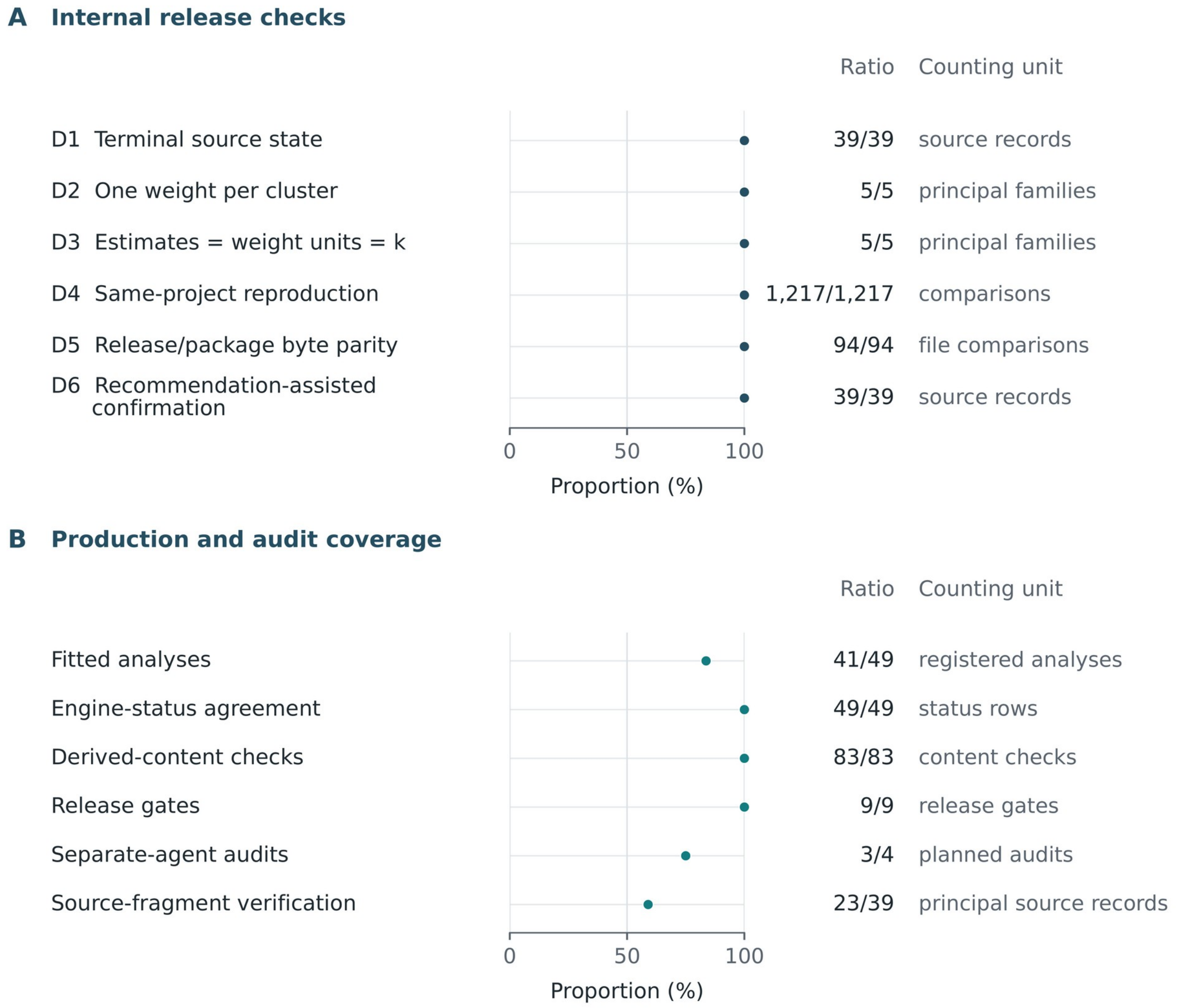


Supplementary Figure S1. Internal release checks and audit coverage. Points show each reported ratio on a common percentage scale. Counting units differ between rows, so the proportions are not an overall accuracy estimate or a combined performance score. (A) Six internal release checks. D1 covers 39 source records from five principal families: 29 source-corrected and 10 locator-verified, with none unresolved. D2 checks one weight per dependence cluster per analysis in the encoded, adjudicated graph without independently validating dependence judgments. D3 checks equality of selected-estimate counts, analysis weight units and reported k in the five principal families. D4 compares two implementations within the project across 41 fitted analyses and 87 quantity types. These comparisons are not an independent validation sample. D5 compares 47 artifacts in two manuscript packages for byte identity, not semantic validity. D6 uses recommendation-assisted two-person confirmation, with item agreement of 312/312 (39 records × 8 questions). Both reviewers saw the same recommendations, so this was not blinded independent review or a gold standard. (B) Production and check completion. Of 49 registered analyses, seven were not fittable ($k < 2$) and one was not poolable. Three of four planned within-project separate-agent audits were completed; source audit A was incomplete. Mechanical source-fragment verification covered 23 principal source records: 15 with normalized source-fragment matches and eight with table-cell token matches. Sixteen principal records were not mechanically verified. k denotes the number of analysis weight units within an analysis.

### Supplementary Figure S2

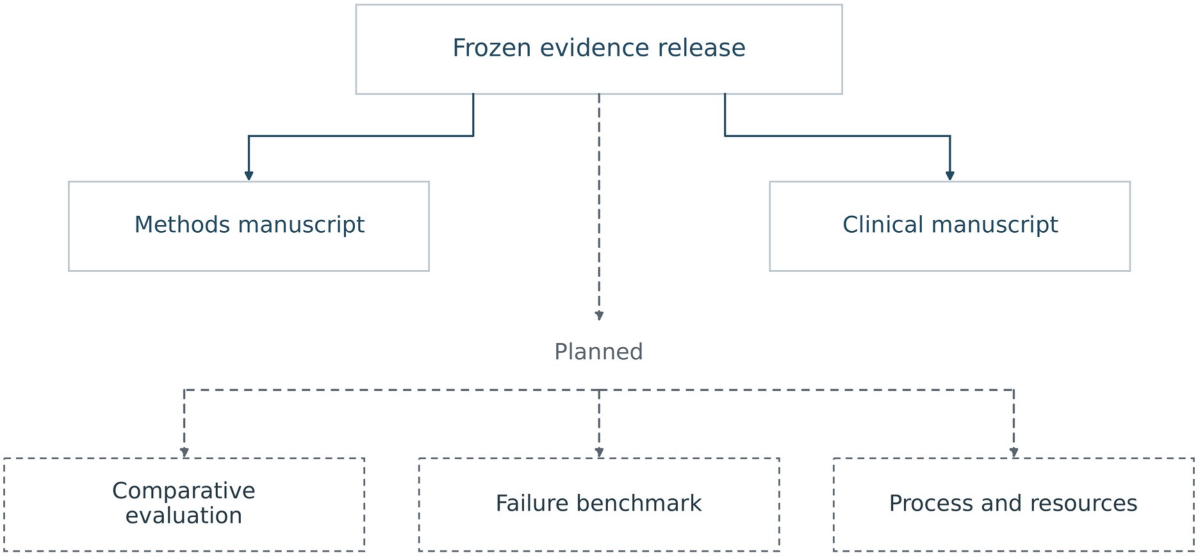


Supplementary Figure S2. Manuscripts and planned evaluations based on frozen evidence release SR-49523c19b885c87a. Solid branches show the methods manuscript, covering historical error pathways, correction consequences and internal release checks, and the clinical manuscript, covering clinical interpretation, forest plots and certainty. Both manuscripts use the same production review; the clinical manuscript is not an independent validation dataset. Dashed branches show planned comparison with an independent human reference, a documented-defect benchmark, and evaluation of workflow and resource use. These planned evaluations were not conducted in this study. The planned benchmark would test failures experimentally, extending the descriptive historical event registry reported here. The manuscripts share scientific values and artifact lineage while addressing different questions.

### Supplementary Figure S3

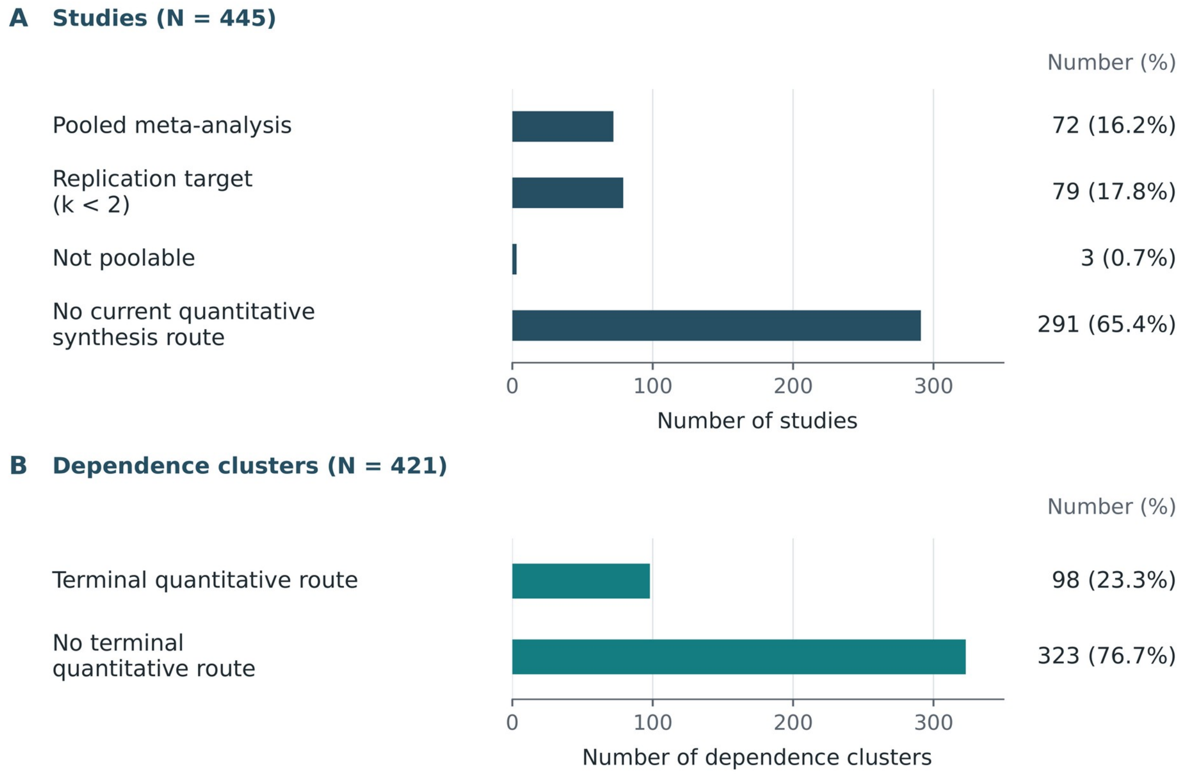


Supplementary Figure S3. Terminal evidence routes. (A) Routes for 445 studies. Three studies were not poolable because their units were incommensurable. (B) Routes for 421 dependence clusters. The 98 clusters with a terminal quantitative route comprise 36 direction-only and 62 effect-size clusters. Percentages use the denominator shown in each panel and may sum to 100.1% because of rounding. Studies and dependence clusters are different counting units. Studies entering pooled meta-analysis are not interchangeable with independent effect estimates. No current quantitative synthesis route does not imply evidence of absence. k denotes the number of analysis weight units within an analysis.

### Appendix S1. AI systems, prompt governance and human oversight

A. Systems and roles. Anthropic, PBC — Claude Code (command-line large-language-model agent): screening, extraction, source-grounded adjudication recommendations, code, orchestration, audit and writing support for the evidence release and for this manuscript. Access: consumer subscription purchased personally by the author; network use: bibliographic services (Crossref, PubMed/Europe PMC, arXiv, OSF), publisher full-text retrieval through the investigator's authenticated institutional browser session, and reading of journal, guideline and vendor web pages for the related-work and journal-requirement checks. Governance: numbered written investigator directives and, once locked, hash-frozen specifications (section D below). OpenAI — ChatGPT (consumer subscription): strategic advice (study planning and scientific positioning); the initial manuscript, figures and review documents were prepared by the author on 2026-08-31. Access: consumer subscription purchased personally by the author. Governance: free-form strategic conversation; no session log retained; model versions and dates of use were not recorded. DeepSeek: not used in the reported workflow. No AI system is listed as an author; the authors retain responsibility for the content. Private or hidden model reasoning traces were not requested and are not disclosed.

B. Model identifiers and dates. Agent execution logs were assessed as of 2026-09-15T18:02:41. The scan covered 52 project sessions across 2 log directories, including 90 top-level and 9330 subagent or workflow transcripts. Identifiers are the strings recorded in the logs; runtime build numbers were not recorded. Assistant messages were counted once per distinct message ID, even when a message spanned several log lines (thinking, text and tool calls), and once overall when forked or resumed sessions repeated earlier messages. Historical production of the shared evidence release and preparation of this manuscript are listed separately. The 5 sessions attributed to this manuscript met the following rule: agent turns in the top-level transcript contained >= 20 lines with an identifier specific to this manuscript. Attached files did not count, and subagent and workflow transcripts inherited the attribution. Assistant messages were counted from the first such line, not from the start of the session. Dates are local calendar dates (UTC+08:00); a session contributes only the messages from its first agent turn that carries an identifier of this manuscript.

| Model identifier | First use | Last use | Assistant messages | Scope |
|---|---|---|---|---|
| claude-fable-5 | 2026-07-13 | 2026-09-03 | 1689 | whole project (shared release production, the companion clinical manuscript and this manuscript) |
| claude-opus-4-8 | 2026-07-13 | 2026-08-13 | 3447 | whole project (shared release production, the companion clinical manuscript and this manuscript) |
| claude-sonnet-5 | 2026-07-22 | 2026-08-11 | 2048 | whole project (shared release production, the companion clinical manuscript and this manuscript) |
| claude-haiku-4-5-20251001 | 2026-07-22 | 2026-07-22 | 60 | whole project (shared release production, the companion clinical manuscript and this manuscript) |
| claude-opus-5 | 2026-07-27 | 2026-09-15 | 92801 | whole project (shared release production, the companion clinical manuscript and this manuscript) |
| claude-fable-5-1 | 2026-09-03 | 2026-09-14 | 3658 | whole project (shared release production, the companion clinical manuscript and this manuscript) |
| claude-opus-5 | 2026-08-31 | 2026-09-15 | 1658 | sessions attributed to this manuscript |
| claude-fable-5 | 2026-09-01 | 2026-09-01 | 8 | sessions attributed to this manuscript |
| claude-fable-5-1 | 2026-09-13 | 2026-09-13 | 361 | sessions attributed to this manuscript |

The overall period was 2026-07-13 to 2026-09-15. Sessions attributed to this manuscript ran from 2026-08-31 to 2026-09-15. The author prepared the initial manuscript, figures and review documents on 2026-08-31. During this earlier period, ChatGPT was used only for strategic advice, and no ChatGPT transcript was retained. Later editorial use is described in section H. The following were not recorded: the Claude Code agent's runtime build identifier; temperature/sampling settings and sampling seeds of the language-model runtime (not exposed by the agent runtime); the selection rule and seed of the risk-weighted full-text verification sample (not archived in the inputs of this study); OpenAI ChatGPT session logs, model versions and dates of use; and the software environment (package versions) of the release build that produced the release statistics and clean-room comparisons.

C. Sequence of phases and dates. This sequence is shared with the companion clinical manuscript because both draw on the same release. Cross-references to its supplement are marked, and people are named by role. Each phase ended with the human action shown. Where individual phase dates were not recorded, a shared date range is given. Frozen artifacts could be changed only by numbered directives. Logs also show agent-assisted work before the first phase: from 2026-07-13, agents helped the investigator draft the review registration and protocol and prepare the two screening prompts. The investigator submitted the registration (PROSPERO CRD420261449181). Agent-assisted title and abstract screening also began before phase 2. From 2026-07-14 to 2026-07-17, two agent reviewers screened 3,905 records from an initial PubMed search, with adjudication of disagreements. On 2026-07-18, the screening plan changed so that all 20,399 deduplicated records, including those initially screened, were screened afresh in phase 2. The earlier decisions served only as a quality-control cross-check.

| Phase | Dates | Standing instruction to the agent | Governing frozen specification | Human act that closed the phase |
|---|---|---|---|---|
| **1. Search and deduplication** | 2026-07-14 to 2026-07-17 | Agent task under investigator direction: agents helped revise the search strategies (including the known-item validation behind the revised strategy), ran the PubMed search through NCBI E-utilities, exported records from several other databases through the investigator's authenticated institutional browser session, and deduplicated the records | Search strategies, reproduced verbatim per database (companion clinical manuscript, eMethods 1) | Last-search date fixed at 2026-07-17; no database searched thereafter (the search-date reconciliation record) |
| **2. Title and abstract screening** | from 2026-07-18 | Judge each record against the eligibility contract and return a coded decision with the clause relied on. Two agents judged independently and blinded to each other; a third adjudicated every disagreement. | The eligibility criteria specification (locked v1.0, 2026-07-29; public deposit, protocol directory) | Investigator rulings on protocol ambiguities, issued as numbered directives |
| **3. Full-text assessment and retrieval** | through 2026-08-05 | Assess each retrieved full text against the same contract; a separate verifier agent, holding the full text, re-judged a risk-weighted sample seeking grounds to overturn the decision. | The eligibility criteria specification (locked v1.0, 2026-07-29; public deposit, protocol directory) | Full-text retrieval closure recorded 2026-08-05 (the frozen PRISMA count record) |
| **4. Corpus closure (identity gap wave)** | 2026-08-04 | Finalise the 28 already-retrieved records still carrying non-terminal states. No new search; within-flow reclassification only. | Corpus closure protocol; PRISMA counts unchanged | PI approval of the material operational clarification, 2026-08-04 |
| **5. Structured extraction** | 2026-07-18 to 2026-08-31 (shared envelope bounded by the first logged session in the review workspace and the signed release; individual phase dates were not recorded) | Extract only values printed in the source into the controlled vocabulary, attaching a verbatim source locator to every analyzed estimate. Never infer, never auto-fill; where a value is not printed, record NOT_REPORTED. | EVIDENCE_REPRESENTATION_CONTRACT_v2; ADJUSTMENT_TWO_AXIS_SPEC_v1 | Third-pass adjudication from source where the dual pass disagreed |
| **6. Dependence adjudication and clustering** | 2026-07-18 to 2026-08-31 (shared envelope bounded by the first logged session in the review workspace and the signed release; individual phase dates were not recorded) | Cluster cohort entities that share participants under the registry-provenance rules; at most one weight unit per cluster per analysis. | COHORT_OVERLAP_RESOLUTION_PROTOCOL_v1 | PI directives established two cluster edges the rules could not settle |
| **7. Estimate selection** | 2026-07-18 to 2026-08-31 (shared envelope bounded by the first logged session in the review workspace and the signed release; individual phase dates were not recorded) | Where several eligible estimates exist for one outcome, select one through the frozen source-based hierarchy. The hierarchy may never use effect direction, interval width or null-crossing; re-run every legal alternative as a multiverse. | ESTIMATE_SELECTION_PROTOCOL; PAPER1_ESTIMATE_SELECTION_PROTOCOL_v1 | PI adjudicated level semantics and ties |

| Phase | Dates | Standing instruction to the agent | Governing frozen specification | Human act that closed the phase |
|---|---|---|---|---|
| **8. Risk of bias and certainty** | 2026-07-18 to 2026-08-31 (shared envelope bounded by the first logged session in the review workspace and the signed release; individual phase dates were not recorded) | Judge QUIPS at the level of the result, not the study; derive result-level judgments from the study level by frozen rule; compute certainty from the frozen GRADE protocol rather than rating de novo. | The formal prognostic GRADE protocol, version 1.1 (public deposit) | PI adjudicated the domain criteria and froze the protocol before application |
| **9. Synthesis** | 2026-07-18 to 2026-08-31 (shared envelope bounded by the first logged session in the review workspace and the signed release; individual phase dates were not recorded) | Fit only what the frozen analysis plan permits; run leave-one-out and the selection multiverse for every principal analysis; never pool across effect-measure types. | Frozen statistical analysis plan (companion clinical manuscript, eMethods 2) | Same-project clean-room statistical reproduction of all locked results |
| **10. Two-person source confirmation** | 2026-08-31 | Agents generated source-grounded recommendations only; the confirmation decisions themselves were not delegated. | FINAL_HUMAN_CONFIRMATION_METHODS | All 39 principal-analysis source records confirmed against the printed sources by Chuan Yin and Zehao Jing, separately; signatures verified against the archived signed document |
| **11. Release authorisation** | 2026-08-31 | Compile the release and run every gate; a build may not be promoted while any gate fails. | the release manifest | PI signature of scientific release SR-49523c19b885c87a |
| **12. Analyzed-size sign-off** | 2026-09-05 | Revisit sizes not established at extraction; one agent re-derived each, a second re-derived it independently; disagreements were escalated, never resolved by the agents. | Analyzed-size protocol (companion clinical manuscript, eMethods 5) | 28 sizes checked against the printed source and signed by 2 investigators (0 corrections); the remaining 18 carry forward as locator-checked cohort totals |
| **13. Manuscript preparation (companion clinical manuscript)** | 2026-08-31 to 2026-09-09 | Draft and revise from the signed release only; zero recomputation; every reported number must resolve to a frozen ledger field. Reference metadata may not be generated by the model. | Signed release SR-49523c19b885c87a | The authors of the companion clinical manuscript reviewed all generated content and accept responsibility for it |
| **14. Preparation of this methods manuscript** | 2026-08-31 to 2026-09-15 | Draft and revise from the signed release only; no scientific value recomputed (section G) | Signed release SR-49523c19b885c87a | The author reviewed all generated content and accepts responsibility for it |

D. What an instruction consisted of. Once the specifications were locked, three layers determined every agent action: (1) a hash-frozen specification (eligibility contract, outcome and exposure vocabularies, estimate-selection hierarchy, dependence-clustering protocol, and risk-of-bias and certainty protocols); (2) a numbered written investigator directive that applied a specification, adjudicated an ambiguity or authorized a build; and (3) the bibliographic record or full text under assessment. For layer 1, the eligibility criteria, certainty protocol, human-confirmation methods and release manifest are already public. Synthesis contracts, controlled vocabularies and schemas are in the deposited materials. The other specifications named in section C are available from the author, as are the layer 2 directives. Layer 3 consists of copyrighted articles identified by bibliographic identifiers and structured source locators. Prompt templates and numbered directives are retained in the project archive and available from the corresponding author. They are not deposited because they quote copyrighted full texts. Prompt templates were frozen by hash and were not varied. Prompt sensitivity was not evaluated. Training-data contamination was not assessed: the appraised reports are published articles that may be present in the training data of the commercial models used.

E. Human oversight. Material ambiguities were escalated on cards recording the options and their measured consequences; the PI decided. Two people confirmed all 39 principal-family source records after seeing the same source-grounded recommendation (not blinded). The release was compiled on 2026-08-30 and signed on 2026-08-31; the first directive for this manuscript was issued on the day of the signature. Tasks never delegated to an agent: authorship decisions; the two-person confirmation decisions; signature of the scientific release. Reference metadata was not generated by a language model; AMA formatting, numbering and citation insertion were performed by agent-written code and checked against a frozen Crossref snapshot and arXiv records.

F. Copyrighted content entered into the models. Bibliographic records exported from licensed databases and the full texts of reports assessed for eligibility were entered into the agent runtime for screening and extraction, obtained through institutional library subscriptions, open-access licences or individual purchase, and used under the respective access terms for eligibility assessment and data extraction only. No copyrighted full text is reproduced in the manuscript, this supplement or the deposited public materials.

G. Manuscript preparation on 13–15 September 2026. Under the author's written instructions, Anthropic Claude Code (model identifiers for these dates are in the table above) was used to regenerate every table, figure and check from the frozen release outputs (R1), verify references against Crossref and arXiv records, and revise the initial manuscript. This included the declarations, related-manuscript status and data-availability statement updated on 15 September 2026. No scientific value was recomputed and no model was rerun on the literature.

H. Manuscript editing and formatting on 16–22 and 28 September and 7 and 9 October 2026. OpenAI ChatGPT with Codex tools assisted with language editing, bilingual alignment, assessment of supplied methodological publications, reference and source-version checks, author-declaration checks, related-work comparisons in Tables S11 and S9, and the clinical-scope mapping in Table S10. The supplement was assembled using the deposited document-construction function and frozen records. Word files were edited with python-docx and checked through LibreOffice rendering. Earlier figures used AI-assisted Graphviz and Matplotlib code. On 18 September, the six final figures were redrawn with AI-assisted Matplotlib code: flow diagrams use single labels; the figure showing selection, evidence identity and analysis participation uses paired count plots, and Figure S1 uses ratio dot plots. Contributor percentages and point positions were calculated from existing frozen counts. Table S5 was divided into two labelled panels retaining all six cases, and table wording was revised for readability. Scientific inputs and statistical results were unchanged. Source definitions and counts were checked against frozen membership and correction records. The interface did not expose model-build or runtime-build identifiers for these editorial sessions. No clinical evidence extraction, statistical synthesis, new model evaluation or clinical validation was performed. The literature comparisons and scope mapping are editorial synthesis. On 20 September, four supplied publications were assessed, two references were added, and the related-work argument and Tables S11 and S9 were updated. Existing figures and frozen scientific results were retained. The authors retain responsibility for the scientific content and final manuscript. On 21 and 22 September 2026, OpenAI Codex (GPT-6; exact model snapshot and runtime build unavailable) was used for language editing, submission-policy checks, document consistency checks and formatting. The supplied Supplementary Figure S1 was inserted into the Word supplement. No scientific inputs, clinical effect estimates or statistical models were changed or rerun. On 28 September 2026, OpenAI Codex assisted with journal-specific shortening, language editing, cross-reference and declaration checks, document formatting, and preparation of the preprint revision. The main manuscript was reframed as system development and internal evaluation. The former main Table 1 and Figure 5 were moved to Supplementary Table S11 and Supplementary Figure S2; references in this supplement were numbered independently. Scientific inputs, clinical effect estimates, statistical models and the frozen release were unchanged. The exact model snapshot and runtime build were unavailable. On 7 October 2026, OpenAI Codex assisted with revision for Systematic Reviews, focusing the narrative on documented evidence-processing errors, their consequences and correction controls. It also assisted with the case-summary table, the reporting applicability map, figure cross-references, declarations, language editing and document formatting. These changes used the existing frozen records and added no study data, independent validation experiment or statistical analysis. In the final 7 October revision, repeated clinical effect estimates were removed from Table S3 while retaining analysis identities, membership checks and links to the companion manuscript and frozen outputs. The terminal-route figure was moved to Supplementary Figure S3. Scientific counts and the frozen release remained unchanged. On 9 October 2026, OpenAI Codex assisted with manuscript and preprint revisions: study-design clarification, event counting, historical correction-record checks, language and formatting. Historical correction steps were distinguished from final clinical estimates and did not form a complete paired series. No new source data, independent evaluation or statistical analysis was added; frozen release values remained unchanged. The exact model snapshot and runtime build were unavailable.

### Supplementary note 1. Status-file discrepancy inside the signed release

The status file (00_STATUS.md) shipped inside both manuscript packages of the signed release states: "ONE PI DECISION IS STILL PENDING: the primary MARCQI report for the readmission family (`PI_BLINDED_CARD_MARCQI_PRIMARY_REPORT.json`). The current build follows the frozen source hierarchy, which prefers S00498; PATCH-02 §3 had specified S00253. This package must NOT be described as fully PI-adjudicated until that card is answered." That decision was made after the status file was written (PI-PATCH-03 §1): in cluster DC-MARCQI-PRIMARY-TKA-READMISSION, study S00498 carries the primary statistical contribution (whole cohort; pre-exposure covariates) and study S00253 is retained as a linked overlapping report providing parallel post-exposure-conditional evidence without weight. The frozen file MARCQI_CLUSTER_ADJUDICATION_FINAL.csv and the PI sign-off (§5) record this final state. The signed release is read-only and was not edited; this note discloses the discrepancy instead of rewriting the signed archive.

### Supplementary note 2. Scope of the present and planned studies

This methods manuscript evaluates system architecture, release integrity, historical failure events, explicit non-synthesis states and governance.

The companion clinical manuscript addresses clinical effect interpretation, certainty, forest plots and surgical implications. It draws on the same frozen release and does not constitute external validation of the system.

Planned future studies include comparisons against a blinded human reference, a failure benchmark with mutation tests and negative controls, and detailed comparisons of time, token use, cost and productivity. They contribute no empirical results to this manuscript.

### Supplementary references

These S-prefixed references support the supplementary material and are numbered independently of the main manuscript.


S1. Farotimi O, Dunn A, Van Lissa CJ, Polanin JR, Mavridis D, Pigott TD. Guidance for manuscript submissions testing the use of generative AI for systematic review and meta-analysis. Res Synth Methods. 2026;17(2):237-239. doi:10.1017/rsm.2025.10058

S2. Wang Z, Cao L, Danek B, Jin Q, Lu Z, Sun J. Accelerating clinical evidence synthesis with large language models. NPJ Digit Med. 2025;8(1):509. doi:10.1038/s41746-025-01840-7

S3. Xu W, Zhang W, Ling F, et al. Manalyzer: end-to-end automated meta-analysis with multi-agent system. arXiv. Preprint posted May 22, 2025; revised January 21, 2026. arXiv:2505.20310. doi:10.48550/arXiv.2505.20310

S4. Taherinezhad M, Maier S, Vitagliano G, Pierri F, Feuerriegel S. AutoSynthesis: an agentic system for automated meta-analysis. arXiv. Preprint posted July 16, 2026. arXiv:2607.15247. doi:10.48550/arXiv.2607.15247

S5. Cao C, Arora R, Cento P, et al. Automation of systematic reviews with large language models. medRxiv. Preprint posted June 13, 2025; version 3 revised February 18, 2026. doi:10.1101/2025.06.13.25329541

S6. Chen SF, Alyakin A, Seas A, et al. LLM-assisted systematic review of large language models in clinical medicine. Nat Med. 2026;32(3):1152-1159. doi:10.1038/s41591-026-04229-5

S7. Li L, Mathrani A, Susnjak T. What level of automation is “good enough”? A benchmark of large language models for meta-analysis data extraction. Res Synth Methods. 2026;17:671-692. doi:10.1017/rsm.2025.10066

S8. Ahuja N, Mulla S, Khan MA, et al. EviSearch: a human in the loop system for extracting and auditing clinical evidence for systematic reviews. arXiv. Preprint, version 2, April 21, 2026. doi:10.48550/arXiv.2604.14165

S9. Markovic M, Indukuri G, Sripada S, et al. Authoring and management of transparent research integrity assessments of randomised clinical trial publications using LLM-assisted tools and provenance knowledge graphs. arXiv. Preprint, version 1, August 7, 2026. doi:10.48550/arXiv.2608.07202

S10. Miao J, Davis JR, Zhang Y, Pritchard JK, Zou J. Reimagining research papers as interactive and reliable AI agents. Nature. Published online September 16, 2026. doi:10.1038/s41586-026-11044-y

S11. Moghaddam MT, Alipour M. ARISMA: guidelines for AI- and LLM-assisted systematic reviews, scoping reviews, and mapping studies. arXiv. Preprint, version 1, August 25, 2026. doi:10.48550/arXiv.2608.25050

S12. Khraisha Q, Put S, Kappenberg J, Warraitch A, Hadfield K. Can large language models replace humans in systematic reviews? Evaluating GPT-4’s efficacy in screening and extracting data from peer-reviewed and grey literature in multiple languages. Res Synth Methods. 2024;15(4):616-626. doi:10.1002/jrsm.1715

S13. Purewal A, Fautsch K, Klasova J, Hussain N, D’Souza RS. Human versus artificial intelligence: evaluating ChatGPT’s performance in conducting published systematic reviews with meta-analysis in chronic pain research. Reg Anesth Pain Med. 2026;51(4):437-442. doi:10.1136/rapm-2024-106358

S14. Sollini M, Pini C, Lazar A, et al. Human researchers are superior to large language models in writing a medical systematic review in a comparative multitask assessment. Sci Rep. 2026;16(1):173. doi:10.1038/s41598-025-28993-5

S15. Kim JK, Chua ME, Li TG, Rickard M, Lorenzo AJ. Novel AI applications in systematic review: GPT-4 assisted data extraction, analysis, review of bias. BMJ Evid Based Med. 2025;30(5):313-322. doi:10.1136/bmjebm-2024-113066

S16. Pratte M, Thirukumar S, Zhang C, Slessarev M, Basmaji J, Prager R. Can large language models approximate the results of meta-analyses in critical care? A meta-research study. J Crit Care. 2026;92:155358. doi:10.1016/j.jcrc.2025.155358

S17. Zou Y, Kim I, Gao N, Li M, Kim MO, Ge J. Closing the screening gap but not the writing gap: a two-topic evaluation of LLMs for systematic reviews and meta-analyses in hepatology. npj Gut Liver. 2026;3:21. doi:10.1038/s44355-026-00068-w

S18. van de Schoot R, de Bruin J, Schram R, et al. An open source machine learning framework for efficient and transparent systematic reviews. Nat Mach Intell. 2021;3(2):125-133. doi:10.1038/s42256-020-00287-7

S19. de Bruin J, Lombaers P, Kaandorp C, et al. ASReview LAB v.2: open-source text screening with multiple agents and a crowd of experts. Patterns (N Y). 2025;6(7):101318. doi:10.1016/j.patter.2025.101318

S20. Ouzzani M, Hammady H, Fedorowicz Z, Elmagarmid A. Rayyan—a web and mobile app for systematic reviews. Syst Rev. 2016;5(1):210. doi:10.1186/s13643-016-0384-4

S21. Valizadeh A, Moassefi M, Nakhostin-Ansari A, et al. Abstract screening using the automated tool Rayyan: results of effectiveness in three diagnostic test accuracy systematic reviews. BMC Med Res Methodol. 2022;22(1):160. doi:10.1186/s12874-022-01631-8

S22. Gartlehner G, Wagner G, Lux L, et al. Assessing the accuracy of machine-assisted abstract screening with DistillerAI: a user study. Syst Rev. 2019;8(1):277. doi:10.1186/s13643-019-1221-3

S23. Hamel C, Kelly SE, Thavorn K, Rice DB, Wells GA, Hutton B. An evaluation of DistillerSR's machine learning-based prioritization tool for title/abstract screening - impact on reviewer-relevant outcomes. BMC Med Res Methodol. 2020;20(1):256. doi:10.1186/s12874-020-01129-1

S24. Bernard N, Sagawa Y Jr, Bier N, Lihoreau T, Pazart L, Tannou T. Using artificial intelligence for systematic review: the example of Elicit. BMC Med Res Methodol. 2025;25(1):75. doi:10.1186/s12874-025-02528-y

S25. Lagisz M, Mizuno A, Morrison K, et al. Using Elicit AI research assistant for data extraction in systematic reviews: a feasibility study across environmental and life sciences. Res Synth Methods. Published online May 29, 2026:1-18. doi:10.1017/rsm.2026.10080

S26. Bloudek LM, Cichewicz AB, Patel K, Sullivan SD, Kallmes KM. A machine learning and large language model tool for systematic literature reviews of health economic evidence: a validation study. PharmacoEconomics. Published online July 29, 2026. doi:10.1007/s40273-026-01648-7

S27. Thomas J, McDonald S, Noel-Storr A, et al. Machine learning reduced workload with minimal risk of missing studies: development and evaluation of a randomized controlled trial classifier for Cochrane Reviews. J Clin Epidemiol. 2021;133:140-151. doi:10.1016/j.jclinepi.2020.11.003

S28. Reason T, Rawlinson W, Langham J, Gimblett A, Malcolm B, Klijn S. Artificial intelligence to automate health economic modelling: a case study to evaluate the potential application of large language models. Pharmacoecon Open. 2024;8(2):191-203. doi:10.1007/s41669-024-00477-8

S29. Susnjak T. A reproducible optimisation protocol for calibrating prompt-based large language model workflows in evidence synthesis. MethodsX. 2026;17:104041. doi:10.1016/j.mex.2026.104041

S30. Flemyng E, Noel-Storr A, Macura B, et al. Position statement on artificial intelligence (AI) use in evidence synthesis across Cochrane, the Campbell Collaboration, JBI and the Collaboration for Environmental Evidence 2025. Environ Evid. 2025;14(1):20. doi:10.1186/s13750-025-00374-5

S31. Thomas J, Hair K, Noel-Storr A, et al. Responsible use of AI in Evidence Synthesis (RAISE 2026) 1: recommendations for practice. Version 3, draft for consultation and revision, March 13, 2026. Open Science Framework. Accessed September 13, 2026. doi:10.17605/OSF.IO/FWAUD

S32. Gallifant J, Afshar M, Ameen S, et al. The TRIPOD-LLM reporting guideline for studies using large language models. Nat Med. 2025;31(1):60-69. doi:10.1038/s41591-024-03425-5

S33. Holst D, Moenck K, Koch J, Schmedemann O, Schüppstuhl T. Transparent reporting of AI in systematic literature reviews: development of the PRISMA-trAIce checklist. JMIR AI. 2025;4:e80247. doi:10.2196/80247

S34. Moher D, Page M, McKenzie J, Takwoingi Y, Mayo-Wilson E. PRISMA-trAIce: a name without endorsement. JMIR AI. 2026;5:e104210. doi:10.2196/104210

S35. Yao Z, Yu H. Toward reviewable medical evidence synthesis for care delivery. npj Digit Med. Published online September 19, 2026. doi:10.1038/s41746-026-03195-z

S36. Gao Z, Lu W, Zhu W, et al. Retrieval-augmented multi-agent framework for evidence-centric medical reasoning. Sci Rep. Published online September 19, 2026. doi:10.1038/s41598-026-72525-8

S37. Schaekermann M, Palepu A, Rodman A, et al. Prospective evidence for conversational medical AI is hard, but non-negotiable. Nat Med. Published online September 14, 2026. doi:10.1038/s41591-026-04639-5

S38. Cao C, Moher D. Conducting systematic reviews in a day: enter artificial intelligence. J Bone Joint Surg Am. 2026;108(4):262-265. doi:10.2106/JBJS.25.01373

S39. Wall C, Li Stange L, Solba H, Gillespie B, Parks D, Godfrey A. Too many bits: tackling the waste epidemic in digital medicine. NPJ Digit Med. 2026;9:188. doi:10.1038/s41746-026-02402-1
