## Additional file 2: Reporting applicability checklist for "Evidence-processing errors and their correction in an LLM-assisted systematic review: a retrospective methodological case study"

### Additional file 2
### Reporting applicability checklist

Evidence-processing errors and their correction in an LLM-assisted systematic review: a retrospective methodological case study

This checklist maps PRISMA 2020 items to a retrospective methodological evaluation of evidence-processing errors in one completed systematic review. It identifies the information reported, its location and the limits of applying review-reporting items to this study. It does not claim full PRISMA compliance for a new clinical systematic review.

The underlying clinical review is the shared development and production case, not an external validation set. Its complete PRISMA flow is reported in the companion clinical review, Figure 1 (doi:10.64898/2026.09.15.26363083). The present manuscript reports the methodological evaluation; clinical items marked as context or not newly assessed do not claim that the clinical report is reproduced here. Quoted locators identify paragraph openings within the named section. Page numbers refer to the 30-page manuscript accompanying this checklist.

Adapted from the PRISMA 2020 checklist: Page MJ, McKenzie JE, Bossuyt PM, et al. BMJ. 2021;372:n71. doi:10.1136/bmj.n71. The original checklist is available at https://www.prisma-statement.org/prisma-2020-checklist under CC BY 4.0 (https://creativecommons.org/licenses/by/4.0/). Item topics are paraphrased; applicability assessments and responses are specific to this manuscript.

| Item and topic | Applicability | What is reported | Location in the manuscript |
|---|---|---|---|
| 1<br>Title | Adapted | The title identifies evidence-processing errors, their correction and the retrospective methodological case-study design. The present article is not a new clinical systematic review. | Title, p. 1. |
| 2<br>Abstract | Adapted | The structured abstract states the question, retrospective non-blinded design, source records, principal error findings and limitations. Registration is explicitly assigned to the underlying review. | Abstract, pp. 1–2, including Background, Methods, Results, Conclusions and Systematic review registration. |
| 3<br>Rationale | Applicable | The rationale concerns errors that arise between source interpretation, identity handling, statistical synthesis and released files, even when individual extracted fields are accurate. | Background, p. 2, paragraph beginning “Large language models (LLMs) support”. |
| 4<br>Objectives | Applicable | The objectives address which documented errors changed the synthesis, how they were detected and corrected, and which implemented controls linked corrections to released outputs. | Background, p. 4, paragraph beginning “We asked which documented evidence-processing errors”. |

| Item and topic | Applicability | What is reported | Location in the manuscript |
|---|---|---|---|
| 5<br>Eligibility criteria | Adapted | The evaluation uses the same completed review in which the system was developed. Confirmed correction events and verified release-blocking findings were counted under explicit rules. Sixty-three ledger rows yielded 39 events after deduplication and root-cause merging; ten Target 4 findings and one infrastructure event completed the total of 50. No new clinical selection was performed. | Methods, p. 4, paragraph beginning "We conducted a retrospective". Methods, p. 5, paragraph beginning "We retrospectively counted distinct root-cause events". Methods, p. 5, paragraph beginning "The event total came from 63 correction-ledger rows". Additional file 1, Table S6. |
| 6<br>Information sources | Adapted | Evaluation materials were drawn from frozen release SR-49523c19b885c87a and its correction, audit and release records. The database counts describe the pre-existing application case; no new search was conducted for this evaluation. | Methods, p. 4, paragraph beginning "We conducted a retrospective". Methods, p. 5, paragraph beginning "We retrospectively counted distinct root-cause events". Declarations, Availability of data and materials, p. 18. |
| 7<br>Search strategies | Not a new review search | This evaluation did not undertake a separate systematic search. Full database search strategies and the complete clinical PRISMA flow are not reproduced in this methodological report. The flow is reported in Figure 1 of the companion clinical review (doi:10.64898/2026.09.15.26363083). | Methods, p. 4, paragraph beginning "We conducted a retrospective". Registration and related work, p. 20; companion clinical review, Figure 1. |
| 8<br>Selection process | Adapted | The manuscript describes LLM-supported screening and human resolution of material ambiguities. Clinical selection belongs to the shared production case. Two reviewers later confirmed selected records after seeing shared recommendations; this was not blinded independent screening validation. | Methods, p. 5, paragraph beginning "The system comprised 14 modules". Methods, p. 9, paragraph beginning "Chuan Yin and Zehao Jing separately reviewed". Figure 1; Additional file 1, Table S2. |
| 9<br>Data collection process | Adapted | Two isolated extraction passes were followed by third-pass source adjudication. Ordered source-authority states governed corrections. Human confirmation used shared source-grounded recommendations and retained escalation decisions. | Methods, p. 5, paragraph beginning "The system comprised 14 modules". Methods, p. 9, paragraph beginning "Chuan Yin and Zehao Jing separately reviewed". Additional file 1, Tables S1 and S2. |
| 10a<br>Outcome definitions | Adapted | Evaluation outcomes include distinct root-cause events, recorded changes in synthesis and package outputs, terminal source states, weight-unit consistency, numerical reproduction, file identity and recommendation-assisted confirmation. These domains retain separate denominators. | Methods, p. 9, paragraph beginning "Six supporting release-integrity domains were reported separately". Methods, p. 5, paragraph beginning "We retrospectively counted distinct root-cause events". Tables 3 and 4 (pp. 28, 29). |

| Item and topic | Applicability | What is reported | Location in the manuscript |
|---|---|---|---|
| 10b<br>Other data fields | Applicable with adaptation | Recorded fields distinguish reports, studies, cohorts, dependence clusters, analysis weight units and estimates. Source-authority states, unknown values, correction histories and release identifiers retain provenance and ambiguity. | Methods, p. 5, paragraph beginning "The system comprised 14 modules". Methods, p. 7, paragraph beginning "The identity graph distinguished". Methods, p. 8, paragraph beginning "Ten release properties covered". Tables 1 and 2 (pp. 26, 27). |
| 11<br>Study risk-of-bias methods | Clinical-case context | The implemented review used result-level QUIPS assessments. Their representation and correction are reported, but their accuracy was not tested against an independent reference standard. | Methods, p. 8, paragraph beginning "Each study received a terminal route". Discussion, p. 16, paragraph beginning "The primary limitation is that". Additional file 1, Table S1. |
| 12<br>Effect measures | Adapted and clinical context | Clinical effect measures remain separate in the implemented engine. Historical paired point estimates were reported only where both values were explicitly recorded and the effect type could be verified. They are correction-stage snapshots, not new pooled results or an isolated-effect experiment. Supporting numerical checks assess same-project agreement. | Methods, p. 8, paragraph beginning "Each study received a terminal route". Methods, p. 5, paragraph beginning "The event total came from 63 correction-ledger rows". Methods, p. 9, paragraph beginning "Six supporting release-integrity domains were reported separately". Table 3 (p. 28). |
| 13a<br>Eligibility for each synthesis | Clinical-case context | Identity and dependence rules determine which estimates may receive a weight in each analysis. The frozen hierarchy selects admissible estimates without using significance or row order. No new clinical evidence set was selected for this article. | Methods, p. 7, paragraph beginning "The identity graph distinguished". Methods, p. 8, paragraph beginning "Each study received a terminal route". Table 1 (p. 26). |
| 13b<br>Data preparation | Applicable with adaptation | Source-authority precedence prevents lower-authority values from replacing adjudicated or verified values. Unknown fields and non-fitted states remain explicit, and corrections trigger specified downstream regeneration. | Methods, p. 5, paragraph beginning "The system comprised 14 modules". Methods, p. 8, paragraph beginning "Each study received a terminal route". Methods, p. 8, paragraph beginning "Ten release properties covered". |
| 13c<br>Tabulation and display | Applicable with adaptation | The three main figures show the architecture, evidence identities and contributions, and historical error events. Tables distinguish corpus units, evaluation domains and error pathways. Supplementary figures report integrity checks, manuscript relationships and terminal evidence routes. | Figures 1–3 (pp. 6, 7, 11); Tables 1–5 (pp. 26, 27, 28, 29, 30); Additional file 1, Supplementary Figures S1–S3. |

| Item and topic | Applicability | What is reported | Location in the manuscript |
|---|---|---|---|
| 13d Synthesis methods | Clinical context and internal check | The implemented engine used restricted maximum likelihood and modified Hartung–Knapp intervals. A second same-project implementation checked numerical agreement. Clinical effects were not refitted for this retrospective evaluation. | Methods, p. 8, paragraph beginning "Each study received a terminal route". Methods, p. 9, paragraph beginning "Six supporting release-integrity domains were reported separately". Results, p. 13, paragraph beginning "For the supporting internal checks, the second implementation reproduced". |
| 13e Heterogeneity exploration methods | No new clinical investigation | The manuscript describes the implemented prediction-interval and small-study-effect thresholds. It does not report a newly conducted investigation of clinical heterogeneity for the methodological evaluation. | Methods, p. 4, paragraph beginning "We conducted a retrospective". Methods, p. 8, paragraph beginning "Each study received a terminal route". |
| 13f Sensitivity analysis methods | Adapted and clinical context | Admissible estimate alternatives entered a multiverse analysis within the underlying review. Historical correction and release checks are evaluated here; they are not an independent sensitivity validation. FE-21 records the separation of postoperative and all-origin survival. | Methods, p. 7, paragraph beginning "The identity graph distinguished". Results, p. 11, paragraph beginning "Time origin (FE-21)". Table 5 (p. 30). |
| 14 Reporting-bias methods | No new clinical assessment | The engine permitted small-study-effect testing at $k \geq 10$ and retained explicit not-reported codes otherwise. This article does not add a new synthesis-level assessment of missing clinical results or publication bias. | Methods, p. 8, paragraph beginning "Each study received a terminal route". Methods, p. 4, paragraph beginning "We conducted a retrospective". |
| 15 Certainty methods | Clinical-case context | Formal principal-family certainty followed GRADE guidance for prognostic factors. The system preserved different claim types and correction records. These procedures were not independently validated as certainty assessments. | Methods, p. 8, paragraph beginning "Each study received a terminal route". Methods, p. 8, paragraph beginning "Ten release properties covered". Additional file 1, Table S1. |
| 16a Selection results and flow | Adapted and clinical context | The Results distinguish 454 reports, 445 studies, 441 cohort entities and 421 dependence clusters. Figure 2 separates these evidence units from analysis contributions; it is not a complete PRISMA flow. The full clinical selection flow remains in the companion review, Figure 1 (doi:10.64898/2026.09.15.26363083). None of these records is a held-out evaluation sample. | Methods, p. 4, paragraph beginning "We conducted a retrospective". Results, p. 13, paragraph beginning "The corpus comprised 454 reports". Figure 2; Table 1 (p. 26); companion clinical review, Figure 1. |

| Item and topic | Applicability | What is reported | Location in the manuscript |
|---|---|---|---|
| 16b Excluded studies and reasons | No new clinical selection | No new clinical study exclusion set was generated for this evaluation. The complete clinical selection flow is reported in the companion review, Figure 1 (doi:10.64898/2026.09.15.26363083). An itemized list of potentially eligible excluded studies is not reproduced in this methodological article. | Methods, p. 4, paragraph beginning "We conducted a retrospective". Registration and related work, p. 20; companion clinical review, Figure 1. |
| 17 Included-study characteristics | Adapted and clinical context | The manuscript characterizes the production corpus, evidence identities, analysis contributions and evaluated records. It does not reproduce the full individual-study clinical characteristics inventory. | Results, p. 13, paragraph beginning "The corpus comprised 454 reports". Results, p. 13, paragraph beginning "All 39 source records". Table 1 (p. 26); Figure 2 (p. 7). |
| 18 Study risk-of-bias results | Clinical-case context | The reported primary aggregation classified 56 of 81 selected results as high risk and 25 as moderate risk. These are same-case outputs; a complete new study-by-study clinical bias assessment is not presented here. | Results, p. 13, paragraph beginning "All 39 source records". Discussion, p. 16, paragraph beginning "The primary limitation is that". |
| 19 Individual-study effects | No new clinical-results synthesis | Table S3 retains analysis and source identifiers, membership hashes, count checks and release provenance for the principal families. Clinical effect estimates and certainty results are referenced to the companion review and frozen archive. Source and numerical checks here evaluate record integrity and internal consistency. | Results, p. 13, paragraph beginning "All 39 source records". Results, p. 13, paragraph beginning "For the supporting internal checks, the second implementation reproduced". Additional file 1, Table S3; Declarations, Availability of data and materials, p. 18. |
| 20a Synthesis characteristics and bias | Clinical-case context | The report describes membership, dependence handling and synthesis contributions. It does not independently reassess the clinical characteristics and risk of bias of every study entering each synthesis. | Methods, p. 7, paragraph beginning "The identity graph distinguished". Results, p. 13, paragraph beginning "The corpus comprised 454 reports". Results, p. 13, paragraph beginning "All 39 source records". Table 1 (p. 26). |
| 20b Statistical synthesis results | Adapted | Twelve of 50 historical root-cause events changed pooled results before correction. Two recorded correction-stage OR contrasts illustrate the consequences: FE-06, 1.2743 to 1.2763 with k 17 to 16; FE-11, 1.0663 to 1.3709 with k unchanged. These are not final clinical estimates or a complete paired evaluation. Supporting numerical checks matched all 1217 comparisons. | Results, p. 10, paragraph beginning "Fifty distinct root-cause events were documented". Results, p. 12, paragraph beginning "Cohort dependence (FE-06)". Results, p. 12, paragraph beginning "Outcome direction (FE-11)". Tables 4 and 5 (pp. 29, 30). |

| Item and topic | Applicability | What is reported | Location in the manuscript |
|---|---|---|---|
| 20c Heterogeneity investigation results | No new clinical investigation | No new results from a clinical heterogeneity investigation were generated for this article. Error categories describe historical process defects and are not clinical heterogeneity findings. | Methods, p. 4, paragraph beginning “We conducted a retrospective”. Results, p. 10, paragraph beginning “The largest root-cause classes were”. |
| 20d Sensitivity analysis results | Adapted and clinical context | The results describe correction consequences, numerical reproduction and release consistency. FE-21 records the historical primary/sensitivity distinction for survival time origin. This article does not add new clinical sensitivity-analysis estimates. | Results, p. 11, paragraph beginning “Time origin (FE-21)”. Results, p. 13, paragraph beginning “For the supporting internal checks, the second implementation reproduced”. Table 5 (p. 30). |
| 21 Reporting-bias results | No new clinical assessment | No new results on missing clinical findings or publication bias are presented in this methodological evaluation. The implementation description of small-study-effect testing does not constitute a new completed bias assessment. | Methods, p. 4, paragraph beginning “We conducted a retrospective”. Methods, p. 8, paragraph beginning “Each study received a terminal route”. |
| 22 Certainty results | Clinical-case context | The same-case certainty registry contained 56 claims, including five formal prognostic grades. FE-38 changed the risk-of-bias domain but left the already-lowest final GRADE category unchanged. The complete clinical certainty results remain with the companion review and frozen archive; these are not ratings of system effectiveness. | Results, p. 13, paragraph beginning “All 39 source records”. Results, p. 12, paragraph beginning “Model selection and certainty (FE-38)”. Table 5 (p. 30); Declarations, Availability of data and materials, p. 18. |
| 23a Interpretation | Applicable with adaptation | The Discussion interprets the documented error pathways alongside automation, source-auditing and evidence-synthesis studies, and distinguishes implemented functions from independent comparative performance. | Discussion, p. 14, paragraph beginning “Twelve of the 50 documented root-cause events”. Discussion, p. 15, paragraph beginning “These connections complement existing systems”. |
| 23b Evidence limitations | Applicable with adaptation | The development review was also the evaluation case. The report states that the retrospective non-blinded design lacks a held-out domain, external comparator and independent human reference, limiting comparative accuracy and generalizability claims. | Discussion, p. 16, paragraph beginning “The primary limitation is that”. |

| Item and topic | Applicability | What is reported | Location in the manuscript |
|---|---|---|---|
| 23c Process limitations | Applicable with adaptation | Limitations include reconstructed event histories, incomplete source-fragment matching, an unfinished audit, shared recommendations, residual release limitations and missing runtime builds. Event counts cannot estimate incidence, detection sensitivity or errors prevented. | Results, p. 14, paragraph beginning “Both reviewers confirmed all 39 principal records”. Discussion, p. 17, paragraph beginning “The event registry was reconstructed”. Methods, p. 9, paragraph beginning “Chuan Yin and Zehao Jing separately reviewed”. |
| 23d Implications | Applicable with adaptation | Implemented checks are offered as inspectable rules for other review teams. Future prospective comparative evaluation is proposed; reductions in error, reviewer work, cost or patient harm were not measured. | Discussion, p. 15, paragraph beginning “The cases suggest concrete checks”. Discussion, p. 17, paragraph beginning “The next evaluation should freeze”. Conclusions, p. 17. |
| 24a Registration | Adapted | PROSPERO CRD420261449181 registers the underlying prognostic review. The present evaluation was retrospective and has no claimed separate prospective registration; its analysis plan was hashed after the release. | Methods, p. 4, paragraph beginning “We conducted a retrospective”. Abstract, p. 2, Systematic review registration; Registration and related work, p. 20. |
| 24b Protocol access | Adapted | The manuscript identifies the public frozen release and supporting contracts, ledgers, checks and derivation code. These are reproducibility materials for this retrospective evaluation, not a prospectively registered independent evaluation protocol. | Methods, p. 4, paragraph beginning “We conducted a retrospective”. Methods, p. 9, paragraph beginning “Six supporting release-integrity domains were reported separately”. Declarations, Availability of data and materials, p. 18. |
| 24c Amendments | Adapted | Correction, recurrence, supersession and release histories retain their timing and terminal status. The event registry reports operational corrections; it is not presented as a complete account of amendments to the clinical review protocol. | Methods, p. 8, paragraph beginning “Ten release properties covered”. Methods, p. 5, paragraph beginning “We retrospectively counted distinct root-cause events”. Results, p. 13, paragraph beginning “For the supporting internal checks, the second implementation reproduced”. Additional file 1, Tables S5–S7. |
| 25 Support | Applicable | No specific grant was received. The corresponding author paid for commercial AI subscriptions and computing. The acknowledgements section states that it is not applicable. | Declarations, Funding, p. 19 and Acknowledgements, p. 19. |

| Item and topic | Applicability | What is reported | Location in the manuscript |
|---|---|---|---|
| 26<br>Competing interests | Applicable | Chuan Yin developed and owns the Evidence OS software used in the review. The other authors declare no competing interests. | Declarations, Competing interests, p. 19. |
| 27<br>Data, code and materials | Applicable | The public archive contains versioned evaluation materials, registries, contracts, ledgers, release checks, deterministic derivation code and both statistical engines. The statement gives the version DOI and licenses and excludes copyrighted full texts, credentials and signed documents. | Declarations, Availability of data and materials, p. 18; Methods, p. 9, paragraph beginning “Chuan Yin and Zehao Jing separately reviewed”. |
